# Three-dimensional cephalometric landmarking and Frankfort horizontal plane construction: reproducibility of conventional and novel landmarks

**DOI:** 10.1101/2021.08.05.21261623

**Authors:** Gauthier Dot, Frédéric Rafflenbeul, Adeline Kerbrat, Philippe Rouch, Laurent Gajny, Thomas Schouman

## Abstract

In some dentofacial deformity patients, especially patients undergoing surgical ortho-dontic treatments, Computed Tomography (CT) scans are useful to assess complex asymmetry or to plan orthognathic surgery. This assessment would be made easier for orthodontists and sur-geons with a three-dimensional (3D) cephalometric analysis, which would require the localiza-tion of landmarks and the construction of reference planes. The objectives of this study were to assess manual landmarking repeatability and reproducibility (R&R) of a set of 3D landmarks and to evaluate R&R of vertical cephalometric measurements using two Frankfort Horizontal (FH) planes as references for horizontal 3D imaging reorientation. Thirty-three landmarks, di-vided into “conventional”, “foraminal” and “dental”, were manually located twice by 3 experi-enced operators on 20 randomly-selected CT scans of orthognathic surgery patients. R&R confi-dence intervals (CI) of each landmark in the -x, -y and -z directions were computed according to the ISO 5725 standard. These landmarks were then used to construct 2 FH planes: a conventional FH plane (orbitale left, porion right and left) and a newly proposed FH plane (midinternal acous-tic foramen, orbitale right and left). R&R of vertical cephalometric measurements were computed using these 2 FH planes as horizontal references for CT reorientation. Landmarks showing a 95% CI of repeatability and/or reproducibility > 2mm were found exclusively in the “conventional” landmarks group. Vertical measurements showed excellent R&R (95% CI < 1mm) with either FH plane as horizontal reference. However, the 2 FH planes were not found to be parallel (absolute angular difference of 2.41°, SD 1.27°). Overall, “dental” and “foraminal” landmarks were more reliable than the “conventional” landmarks. Despite the poor reliability of the landmarks orbitale and porion, the construction of the conventional FH plane provided a reliable horizontal reference for 3D craniofacial CT scan reorientation.

## Introduction

Diagnosis and planning of orthodontic and maxillofacial treatments rely heavily on X-ray imaging. Twodimensional (2D) X-rays are routinely used but result in a flattening of three-dimensional (3D) craniofacial structures. In some clinical cases of dentofacial deformities – especially patients undergoing surgical orthodontic treatments (orthognathic surgery) – Computed Tomography (CT) or Cone Beam CT (CBCT) scans are useful [1]. For example, 3D imaging makes it possible to assess complex asymmetry and to obtain highly accurate orthognathic surgery planning that can subsequently be used for the manufacturing of surgical guides [1–5]. Several methods have recently been proposed for a fully automatic detection of the best symmetry plane in craniofacial CT scans [6,7]. The diagnostic value of these scans would increase if they could be used to perform 3D cephalometric analysis, which would require the localization of landmarks [8]. At the time being, however, no set of 3D landmarks has been deemed sufficiently reproducible and repeatable for 3D cephalometry [8,9].

Most of the time, three-dimensional cephalometric landmarks previously tested in repeatability and reproducibility studies derived from classic 2D analysis [9]. Some of these landmarks have been shown to be poorly reproducible in 3D, especially orbitale (Or), porion (Po), gonion (Go), condylion (Co) and ramus (Ra) [10–19]. The localization of midsagittal landmarks has generally been shown to be reliable, mostly in datasets of patients showing no asymmetries [8,9]. Several authors suggested using “new” landmarks which cannot be localized on 2D X-rays. More specifically, landmarks located on the craniofacial foramens are presumably easy to identify and should provide good reproducibility [9,14,15]. However, few studies have tested the reproducibility of the new landmarks, and their reliability has not been tested yet in the context of presurgical orthodontic patients [15,20].

The main goal of cephalometric landmarking is to measure distances and angles between landmarks and planes so as to obtain a cephalometric analysis. In order to provide clinically relevant measurements that can be decomposed in the 3 planes of space (i.e. anteroposterior, vertical and transversal), 3D images need to be reoriented in a generic coordinate system [21,22]. The Frankfort Horizontal (FH) plane, used for standardizing and unifying the measurements, is the most commonly used horizontal reference for this coordinate system [21,23]. Its 3D clinical value has been demonstrated for assessing craniofacial morphology and evaluating soft-tissue and skeletal cants in patients receiving orthognathic surgery [24–26]. This plane is conventionally defined in 3D by the 3 following points: left orbitale (Or-L), right porion (Po-R) and left porion (Po-L) [23]. Hence, this reference plane is based on landmarks that are known to be poorly reproducible in 3D, suggesting that the conventionally defined FH plane is poorly reproducible [20]. However, landmark reproducibility does not necessarily result in plane reproducibility, as the latter depends on the direction of landmark errors [14]. To our knowledge, no study has yet tested the repeatability and reproducibility of vertical cephalometric measurements using the conventional FH plane (constructed from 3 landmarks) as a reference for horizontal head reorientation.

Looking for a new plane which would remain parallel with the conventional FH plane but be based on more reliable landmarks, Pittayapat *et al*. suggested a novel FH plane, in which the internal acoustic foramina (IAF) would replace Po [[20]]. Results from experiments performed on CBCT scans of dry human skulls revealed that the localization of IAF provided better reproducibility than that of Po. Moreover, the authors suggested that another new FH plane, based on mid-IAF, Or-R and Or-L, might replace the conventional FH plane, the angular difference found between the two planes being inferior to 1 degree. These results have not been validated yet on 3D scans of living human subjects.

In this context, using a dataset of preoperative CT scans, the aims of our study were:

1. to assess landmarking repeatability and reproducibility of a set of 33 landmarks containing “conventional”, “foraminal” and “dental” landmarks;
2. to assess repeatability and reproducibility of vertical cephalometric measurements using either the conventional or the newly proposed FH planes as references for horizontal head reorientation;
3. to assess the parallelism between the conventional and the newly proposed FH planes.

## Subjects and Methods

### Dataset

Sample size calculation was performed in order to ensure an uncertainty in the repeatability or reproducibility result of 15% for 6 repetitions [27]. As a result, a sample of at least 17 subjects was needed for this study. We performed a random selection of 20 CT scans (7 males, 13 females, mean age 25 ± 8 years) in a database of 134 consecutive orthognathic surgery patients (49 males, 85 females, mean age 27 ± 10 years) from a single Maxillofacial Surgery Department. Patients were considered for inclusion whatever maxillomandibular deformity they presented, with no minimum age. Exclusion criteria were refusal to participate in the research (all patients were contacted by mail) and lack of CT scan segmentation. We used a random number generator to obtain a random sequence of 20 numbers, which was used to select the sample of CT scans included in this study. Allocation was performed by one operator (#1) and supervised by a second operator (#2) at the beginning of the study. All selected subjects showed marked skeletal deformities: 14 skeletal class II - prognathic maxilla and/or retrognathic mandible - (10 short faces, 4 long faces) and 6 skeletal class III - retrognathic maxilla and/or prognathic mandible - (2 short faces, 4 long faces). Six subjects exhibited mandibular asymmetry (2 severe, 4 slight) and 2 subjects exhibited syndromic or rare dentofacial deformities (cleidocranial dysplasia and oligodontia, respectively). A set of 5 random CT scans not included in this study was used for operator training prior to landmarking.

The 20 CT scans were acquired on a Discovery CT750 HD scanner (GE Healthcare, Chicago, USA) set at 100kVp, 50mAs, exposure time 730ms, slice thickness 0.625mm and slice increment 0.320mm. Field of view ranged from 200 to 267mm and pixel size ranged from 0.39 to 0.52mm. Scans were not reoriented after their acquisition. Segmentation of the bones (upper skull, mandible) and upper/lower teeth was performed prior to the study according to an industry-certified semi-automatic process (Materialise, Leuven, Belgium). The study was approved by an Institutional Review Board (IRB No. CRM-2001-051), and all experiments were performed in accordance with relevant guidelines and regulations. Informed consent was obtained from all subjects and/or their legal guardian(s) for participants below age 16 years (all patients were contacted by mail).

### Landmark annotation

The 33 landmarks (Figure 1) were divided into 3 groups: “conventional” (Table 1) “foraminal” (Table 2) and “dental” (Table 3). Operators #1, #2 and #3 (2 trained orthodontists with at least 5 years of clinical experience, 1 final year postgraduate maxillofacial surgeon) received written and verbal instructions on the 3D description and annotation procedure for each landmark (Supplementary Material 1). Manual reorientation of the CT scans was performed based on the Frankfort Horizontal plane construction obtained from the annotation process. A calibration session was organized before the study began, and the instructions were repeated to the operators once more before the second annotation session.

**Figure 1:**
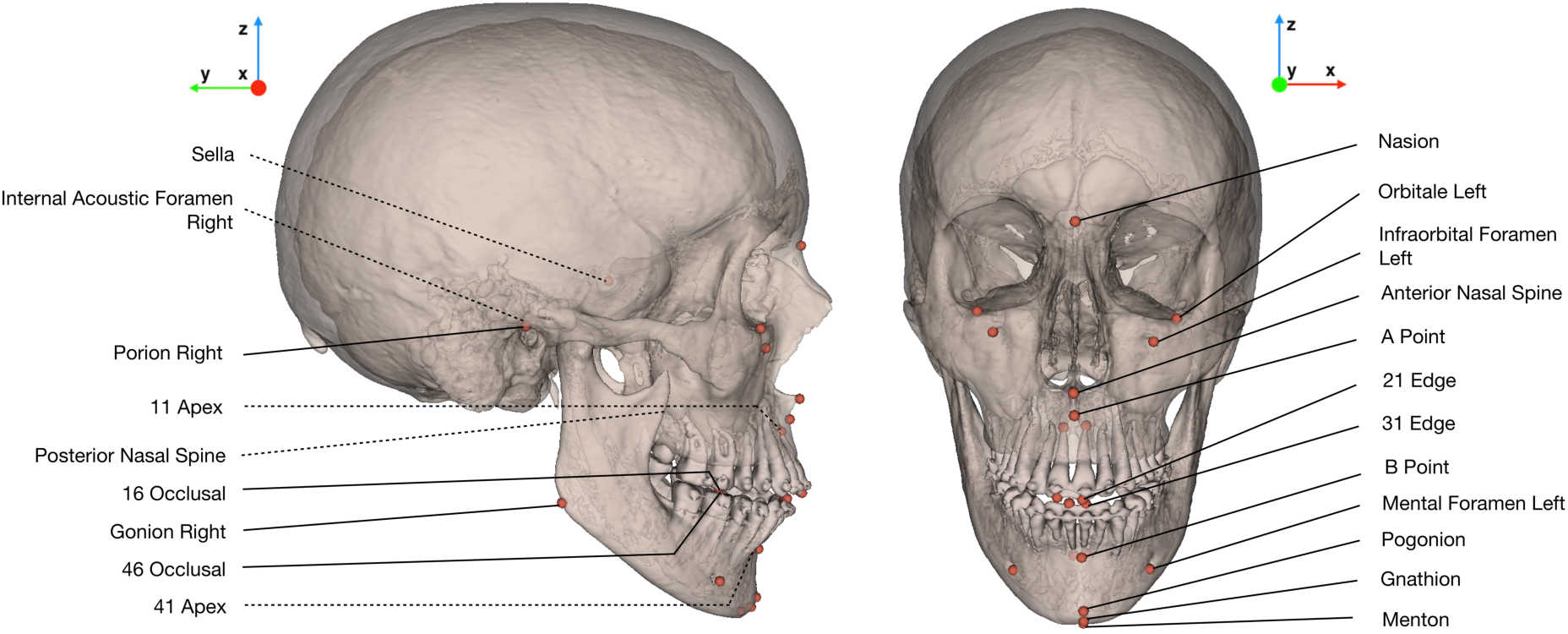
Illustration of the set of 33 landmarks localized by the operators, and the new coordinate system used for statistical analysis. In the case of bilateral landmarks, only one of the two landmarks is labelled. Dotted lines show landmarks localized inside bony structures.

**Table 1:** Definition of “conventional” landmarks localized in our study (L/R: Left/Right)

| Landmark name | Description |
| --- | --- |
| Nasion (Na) | Medial (and upper) point of the frontonasal suture |
| Orbitale L/R (Or-L / Or-R) | Lowest point of the orbital rim L/R |
| Anterior Nasal Spine (ANS) | Medial and most anterior point of the nasal spine |
| A Point (A) | Medial and most posterior point of the maxilla |
| B Point (B) | Medial and most posterior point of the mandible |
| Pogonion (Pog) | Medial and most anterior point of the mandible |
| Gnathion (Gn) | Medial and midpoint between Pog and Me |
| Menton (Me) | Medial and lowest point of the mandible |
| Gonion L/R (Go-L / Go-R) | Midpoint of the gonial angle L/R |
| Porion L/R (Po-L / Po-R) | External & uppermost point of the auditory canal L/R |
| Posterior Nasal Spine (PNS) | Medial & most distal point of the osseous palate |
| Sella (S) | Central point of the sella |

**Table 2:** Definition of “foraminal” landmarks localized in our study (L/R: Left/Right)

| Landmark name | Description |
| --- | --- |
| Infraorbital Foramen L/R (IF-L / IF-R) | External & most distal point of the infraorbital foramen L/R |
| Mental Foramen L/R (MF-L / MF-R) | External & most mesial point of the mental foramen L/R |
| Internal Acoustic Foramen L/R (IAF-L / IAF-R) | External, most mesial and posterior point of the internal acoustic foramen L/R |

**Table 3:** Definition of “dental” landmarks localized in our study (FDI World Dental Federation notation for teeth numbering)

| Landmark name | Description |
| --- | --- |
| 11, 21, 31, 41 edges (11E, 21E, 31E, 41E) | Midpoint of 11/21/31/41 incisal edges |
| 11, 21, 31, 41 apexes (11A, 21A, 31A, 41A) | Root apex of 11/21/31/41 |
| 16, 26 occlusal (16O, 26O) | Summit of the mesiopalatal cusp of 16/26 |
| 36, 46 occlusal (36O, 46O) | Central fossa of 36/46 |

The 20 CT scans and their segmentations were handed over to the 3 operators without any annotations. Manual placement of the 33 landmarks was performed independently by the operators on the software Mimics (v.22.0, Materialise, Leuven, Belgium), and was repeated once after a 3-week interval. Landmarks could be annotated either on the 3D surface or in the Multi-Planar Reconstruction (MPR) views. The operators had neither access to each other’s results nor to their first session’s results when performing the second session. For each session and each CT scan, results were exported as an .xml file containing the x-, y-, z-coordinates of each landmark. The time needed for each CT scan annotation was recorded by the operators and exported in a spreadsheet.

### Statistical analysis

#### New coordinate system for each CT scan

After the two annotation sessions, each CT scan was reoriented in a new coordinate system according to the mean Frankfort Horizontal plane resulting from the 6 repetitions (mean Po-R, mean Po-L, mean Or-L). The origin was set at mid-porion; the -x axis followed the sagittal plane (from right to left); the - y axis followed the frontal plane (from front to back); and the -z axis followed the axial plane (from toe to head) (Figure 1). The landmarking results were then referenced in the new coordinate system before performing the statistical analysis.

#### Landmark repeatability and reproducibility

For each landmark, repeatability and reproducibility standard deviations (SD) were computed according to the ISO 5725 standard of the International Organization for Standardization [28]. Upon initial inspection of the results, the standard’s recommendations were followed for clear outlier points, whose annotations were considered as missing data. The reliability of each landmark in the -x, -y and -z directions was then estimated, considering a 95% confidence interval (CI) of 2*SD of repeatability and reproducibility. Modified Bland-Altman plots, showing the deviations of the landmark positions from their means for the 20 CT scans, were computed for each landmark and direction [29,30].

#### Repeatability and reproducibility of vertical measurements with the conventional FH plane and the newly proposed FH plane

For each CT scan and landmarking session (3 operators, 2 repetitions), we computed the landmarks’ orthogonal projections on 2 FH planes: the conventional FH plane (Or-L, Po-R, Po-L) and the newly proposed FH plane (Or-R, Or-L, mid-IAF). The results were used to compute the standard deviations of repeatability and reproducibility (ISO 5725 standard) of the landmarks’ vertical measurements, using the 2 FH planes as horizontal reference.

#### Parallelism between conventional and newly proposed FH planes

In order to assess whether the conventional FH plane and the new FH plane were parallel, the orthogonal projections of points IAF-R and IAF-L were computed on the mean conventional FH plane (as defined previously) for each subject. We then computed the absolute angular differences between the conventional FH plane and the novel FH plane, using trigonometry to calculate the angles between the normals to the planes.

#### Time needed for landmark localization

Mean time needed and standard deviation for landmark localization were computed.

All data were analysed using the softwares Matlab (v.R2020a, MathWorks, Natick, MA, USA) and RStudio (v.1.3, RStudio PBC, Boston, MA, USA).

## Results

### Landmark repeatability and reproducibility

Outliers were identified for mental foramen points right/left placed by operator #3 during the first annotation session (subjects 4 to 20) and were considered as missing data (Supplementary Materials 2). Repeatability and reproducibility results for the 33 landmarks are shown in Table 4. The landmarks with 95% CI of repeatability and/or reproducibility superior to 2mm for one of their axes were exclusively found in the “conventional” landmark group: point B (-z axis), gonion right/left (-y and –z axes), orbitale right/left (-x axis) and porion right/left (-x axis). Figure 2 shows an example of the modified Bland-Altman plots obtained for five left landmarks: three “foraminal” landmarks (IAF-L, infraorbital foramen left (IF-L), mental foramen left (MF-L)) and two “conventional” landmarks (Or-L and Po-L).

**Table 4:** 95% confidence interval (2*SD) of repeatability and reproducibility of the landmarks (mm), following the ISO 5725 standard. Values between 1 and 2mm are highlighted in orange, and values superior to 2mm are highlighted in red. Repet., repeatability; Repro., reproducibility; SD, standard deviation; L/R: Left/Right.

| Landmark | X Axis |  | Y Axis |  | Z Axis |  |
| --- | --- | --- | --- | --- | --- | --- |
|  | Repet.<br>2*SD | Repro.<br>2*SD | Repet.<br>2*SD | Repro.<br>2*SD | Repet.<br>2*SD | Repro.<br>2*SD |
| **11 Apex** | 0.24 | 0.35 | 0.38 | 0.58 | 0.28 | 0.38 |
| **11 Edge** | 0.25 | 0.34 | 0.24 | 0.30 | 0.08 | 0.10 |
| **16 Occlusal** | 0.63 | 0.76 | 1.27 | 1.51 | 0.21 | 0.28 |
| **21 Apex** | 0.30 | 0.41 | 0.33 | 0.58 | 0.28 | 0.47 |
| **21 Edge** | 0.29 | 0.39 | 0.26 | 0.34 | 0.05 | 0.08 |
| **26 Occlusal** | 0.65 | 0.83 | 0.68 | 0.88 | 0.17 | 0.23 |
| **31 Apex** | 0.18 | 0.24 | 0.25 | 0.32 | 0.27 | 0.36 |
| **31 Edge** | 0.40 | 0.26 | 0.18 | 0.25 | 0.14 | 0.10 |
| **36 Occlusal** | 0.63 | 0.87 | 1.15 | 1.43 | 0.38 | 0.47 |
| **41 Apex** | 0.18 | 0.22 | 0.35 | 0.50 | 0.23 | 0.35 |
| **41 Edge** | 0.19 | 0.24 | 0.20 | 0.22 | 0.05 | 0.07 |
| **46 Occlusal** | 0.40 | 0.54 | 0.55 | 0.82 | 0.23 | 0.30 |
| **A Point** | 0.80 | 0.86 | 0.29 | 0.34 | 1.31 | 1.59 |
| **Anterior Nasal Spine** | 0.57 | 0.67 | 0.80 | 1.31 | 0.57 | 1.10 |
| **B Point** | 0.65 | 1.12 | 0.55 | 0.63 | 2.46 | 2.89 |
| **Gnathion** | 0.75 | 1.14 | 0.76 | 0.92 | 1.05 | 1.25 |
| **Gonion L** | 0.63 | 0.93 | 1.55 | 2.02 | 1.96 | 2.64 |
| **Gonion R** | 0.61 | 0.82 | 1.38 | 1.75 | 1.94 | 2.45 |
| **Infraorbital Foramen L** | 0.88 | 0.93 | 0.81 | 1.01 | 0.63 | 0.93 |
| **Infraorbital Foramen R** | 0.87 | 0.99 | 0.80 | 0.93 | 0.66 | 0.82 |
| **Internal Acoustic Foramen L** | 0.52 | 0.79 | 0.81 | 1.09 | 0.84 | 1.15 |
| **Internal Acoustic Foramen R** | 0.51 | 0.82 | 0.88 | 1.04 | 0.56 | 1.01 |
| **Mental Foramen L** | 0.23 | 0.38 | 0.26 | 0.47 | 0.30 | 0.47 |
| **Mental Foramen R** | 0.23 | 0.35 | 0.25 | 0.45 | 0.32 | 0.43 |
| **Menton** | 0.76 | 1.11 | 1.29 | 1.84 | 0.37 | 0.54 |
| **Nasion** | 0.41 | 0.52 | 0.22 | 0.27 | 0.62 | 0.84 |
| **Orbitale L** | 2.05 | 3.40 | 1.00 | 1.82 | 0.40 | 0.56 |
| **Orbitale R** | 1.83 | 3.23 | 1.07 | 1.75 | 0.37 | 0.46 |
| **Posterior Nasal Spine** | 0.29 | 0.53 | 0.36 | 0.50 | 0.61 | 0.92 |
| **Pogonion** | 0.71 | 1.15 | 0.38 | 0.49 | 1.64 | 1.99 |
| **Porion L** | 2.39 | 2.84 | 0.88 | 1.28 | 0.61 | 0.71 |
| **Porion R** | 2.16 | 2.73 | 1.13 | 1.37 | 0.69 | 0.87 |
| **Sella** | 0.94 | 1.12 | 0.60 | 0.70 | 0.82 | 1.15 |

**Figure 2:**
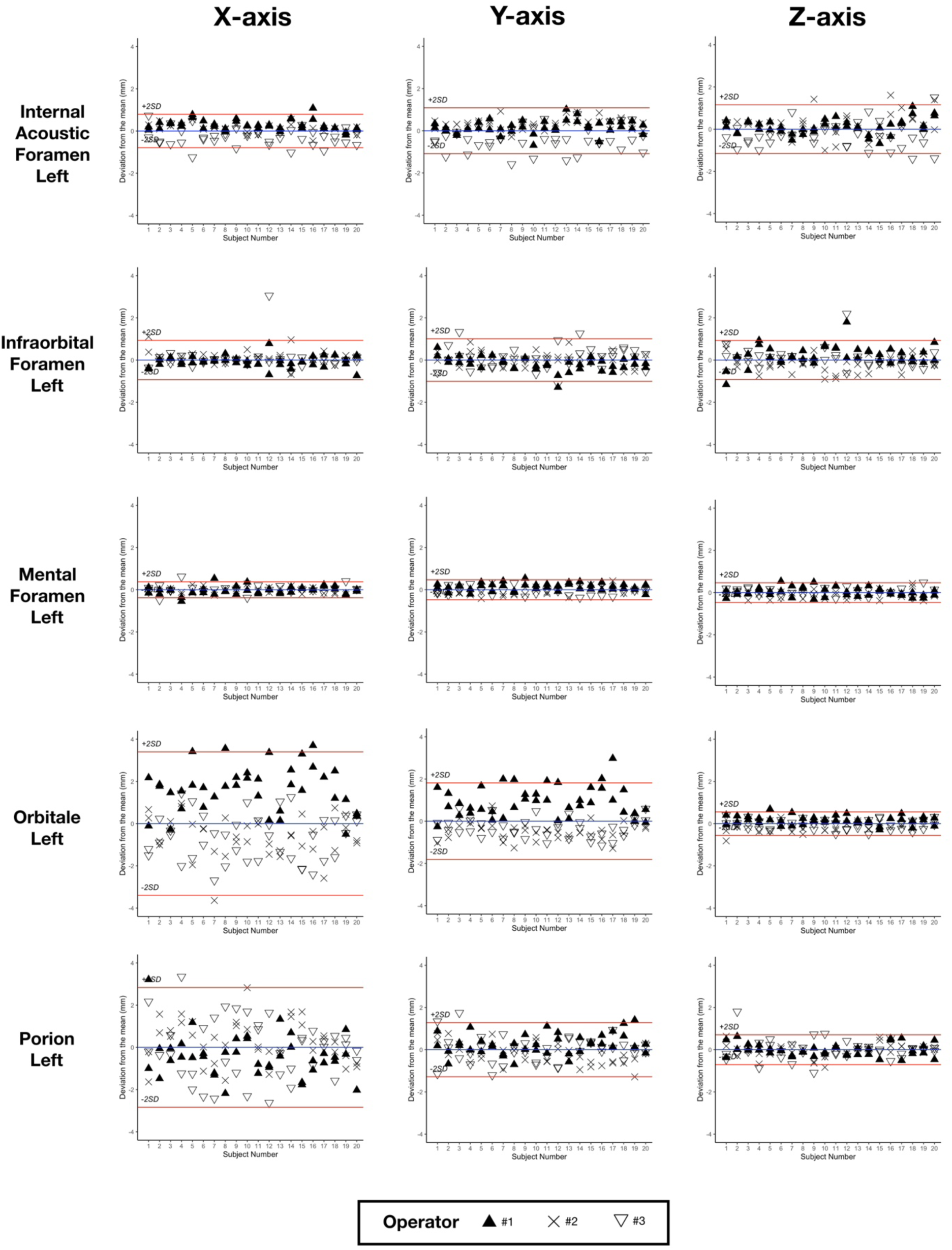
Bland-Altman plots for five left landmarks, showing the deviations from the mean (blue line) of the 6 repetitions for the 20 subjects. Red lines show the ± 2*SD of reproducibility. SD, standard deviation.

### Repeatability and reproducibility of conventional and newly proposed FH planes

The results of the repeatability and reproducibility analysis of vertical measurements of the landmarks using the 2 different FH planes as horizontal reference are shown in Table 5.

**Table 5:** 95% confidence interval (2*SD) of repeatability and reproducibility of the vertical measurements of the landmarks (mm) using 2 FH planes as horizontal references, following the ISO 5725 standard. FH, Frankfort Horizontal plane; Repet., repeatability; Repro., reproducibility; SD, standard deviation; L/R: Left/Right.

| Landmark | Conventional FH |  | Novel FH |  |
| --- | --- | --- | --- | --- |
|  | Repet. 2*SD | Repro. 2*SD | Repet. 2*SD | Repro. 2*SD |
| **11 Apex** | 0.54 | 0.74 | 0.35 | 0.46 |
| **11 Edge** | 0.61 | 0.82 | 0.40 | 0.54 |
| **16 Occlusal** | 0.51 | 0.68 | 0.29 | 0.39 |
| **21 Apex** | 0.53 | 0.71 | 0.35 | 0.46 |
| **21 Edge** | 0.58 | 0.78 | 0.40 | 0.54 |
| **26 Occlusal** | 0.37 | 0.54 | 0.31 | 0.43 |
| **31 Apex** | 0.52 | 0.68 | 0.32 | 0.42 |
| **31 Edge** | 0.57 | 0.75 | 0.37 | 0.49 |
| **36 Occlusal** | 0.35 | 0.52 | 0.31 | 0.43 |
| **41 Apex** | 0.52 | 0.69 | 0.31 | 0.42 |
| **41 Edge** | 0.58 | 0.77 | 0.37 | 0.49 |
| **46 Occlusal** | 0.47 | 0.64 | 0.29 | 0.40 |
| **A Point** | 0.58 | 0.77 | 0.37 | 0.49 |
| **Anterior Nasal Spine** | 0.61 | 0.81 | 0.39 | 0.53 |
| **B Point** | 0.53 | 0.70 | 0.33 | 0.45 |
| **Gnathion** | 0.53 | 0.70 | 0.33 | 0.45 |
| **Gonion L** | 0.48 | 0.56 | 0.52 | 0.74 |
| **Gonion R** | 0.59 | 0.76 | 0.46 | 0.70 |
| **Infraorbital Foramen L** | 0.41 | 0.58 | 0.35 | 0.48 |
| **Infraorbital Foramen R** | 0.62 | 0.80 | 0.33 | 0.43 |
| **Internal Acoustic Foramen L** | 0.60 | 0.71 | 0.61 | 0.91 |
| **Internal Acoustic Foramen R** | 0.63 | 0.80 | 0.59 | 0.90 |
| **Mental Foramen L** | 0.40 | 0.55 | 0.32 | 0.43 |
| **Mental Foramen R** | 0.55 | 0.72 | 0.30 | 0.39 |
| **Menton** | 0.50 | 0.67 | 0.31 | 0.42 |
| **Nasion** | 0.56 | 0.75 | 0.35 | 0.46 |
| **Orbitale L** | 0.40 | 0.56 | 0.38 | 0.50 |
| **Orbitale R** | 0.65 | 0.84 | 0.34 | 0.43 |
| **Posterior Nasal Spine** | 0.39 | 0.53 | 0.31 | 0.46 |
| **Pogonion** | 0.54 | 0.71 | 0.33 | 0.46 |
| **Porion L** | 0.61 | 0.71 | 0.65 | 0.93 |
| **Porion R** | 0.69 | 0.87 | 0.59 | 0.88 |
| **Sella** | 0.43 | 0.55 | 0.40 | 0.60 |

### Parallelism between conventional and novel FH planes

When using the mean conventional FH plane as horizontal reference, the mean absolute vertical measurements ± SD of IAF-L and IAF-R were 2.68 ± 2.51mm and 2.78 ± 2.29mm, respectively. Measurement results for each subject and each repetition are shown in Figure 3. The absolute angular difference between the conventional and the novel FH planes was 2.41° (SD 1.27°).

**Figure 3:**
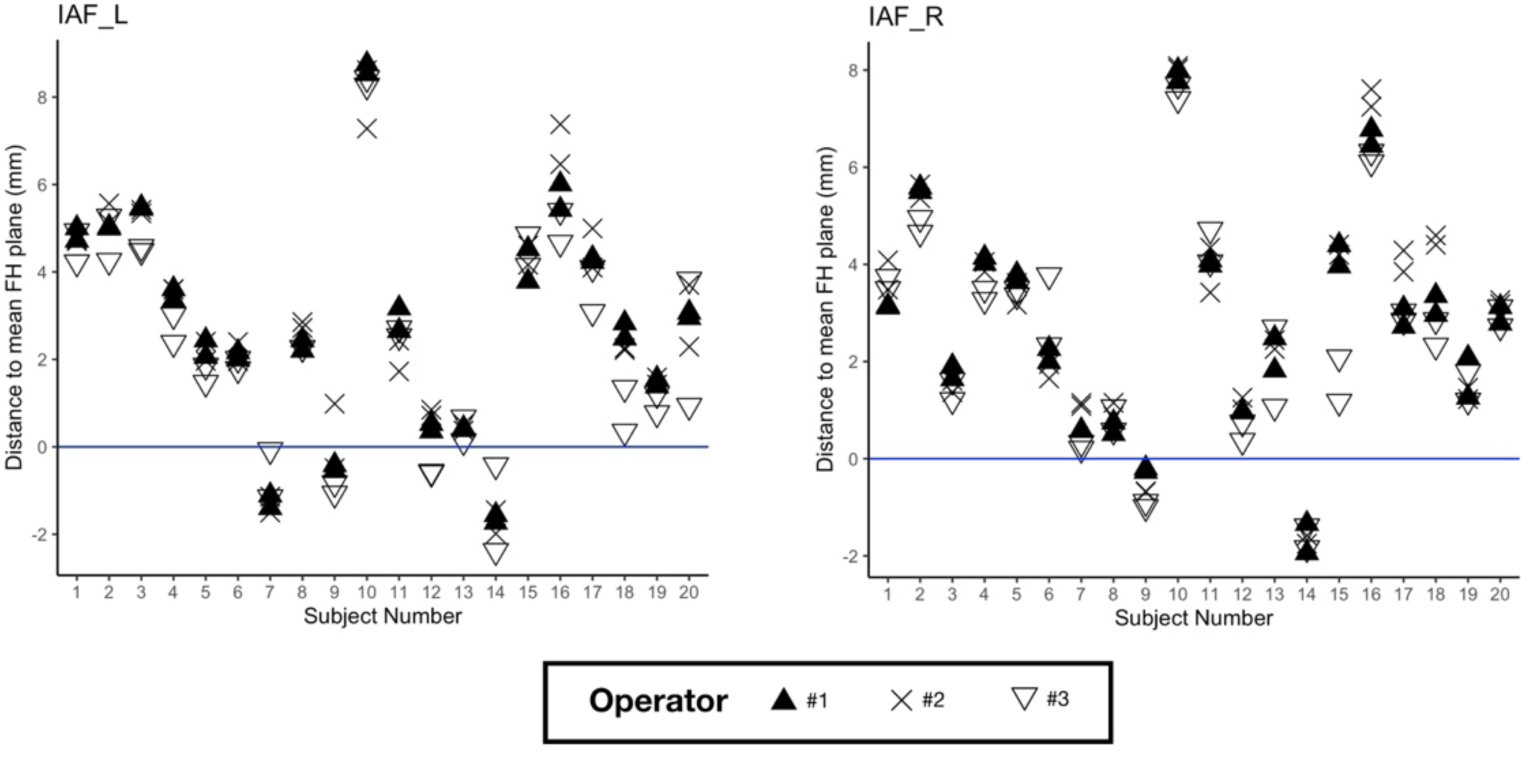
Vertical measurements of IAF left (on the left) and right (on the right) for each subject and repetition, using the mean conventional FH plane as horizontal reference.

### Time needed for landmark localization

The average time required to landmark one CT scan was 14:48 ± 03:45 minutes.

## Discussion

The reliability of 3D cephalometric landmarking and Frankfort Horizontal plane construction is a recurrent clinical issue in orthodontics and orthognathic surgery planning. In this study, we performed a repeatability and reproducibility analysis of conventional and 3D-specific cephalometric landmarks using a database of 20 randomly selected routine presurgical CT scans.

The first aim of our study was to assess landmarking reliability in a set of 33 landmarks containing “conventional”, “foraminal” and “dental” landmarks. As in previously published studies, we ranked the landmarks based on the 95% CI results: landmark with clinically acceptable error when the 95% CI was below 1mm; landmark useful in most analyses when the 95% CI was between 1 and 2mm (highlighted in orange in Table 4); landmark to be used with caution when the 95% CI was above 2mm (highlighted in red in Table 4) [14,16]. Using this classification, all “dental” and “foraminal” landmarks showed a clinically acceptable error or were considered useful in most analyses (16O, 36O, IF-L, IAF-L, IAF-R). The group of “conventional” landmarks showed several landmarks to use with caution (B point, gonion right and left (Go-R, Go-L), Or-R, Or-L, Po-R, Po-L). These findings are in line with previously published reproducibility studies, in which “conventional” landmarks resulting from 2D cephalometric analysis are subject to caution [10–19]. As shown in the Bland-Altman plots (Figure 2), “foraminal” landmarks IAF-L, IF-L and MF-L lead to better repeatability and reproducibility than “conventional” landmarks Or-L and Po-L. We chose not to perform statistical tests such as intraclass correlation coefficients or paired t-tests because of their proven inadequacy in measuring landmarking reliability [29,30]. Outliers were only found in mental foramen landmarks. The definition of this specific “new” landmark was initially ill-understood by one of the operators, who located the landmark at the distal end of the foramen instead of the mesial end, as was agreed upon (Supplementary Materials 2). This illustrates the challenges encountered with landmark identification, which requires very precise definitions in order to be reliable [15]. The fact that our dataset was made of presurgical CT scans did not appear to influence the results negatively when compared with non-surgical subjects in the literature [10,11,13– 18].

Our second objective was to assess the repeatability and reproducibility of vertical 3D cephalometric measurements using either the conventional FH plane or the newly proposed FH plane as reference for horizontal head reorientation. The measurements showed excellent repeatability and reproducibility (95% CI<1mm), using either FH plane as reference. These results tend to prove that despite the poor reliability of the landmarks used to construct them, both planes can be used as reliable horizontal references for head reorientation. An explanation could be that the poor reproducibility of Or, Po and IAF points mainly concerns the -x and -y coordinates. Our results show that a simple method using only 3 landmarks can provide a reliable horizontal reference for 3D head reorientation. Other methods, such as manual reorientation of the 3D model along the FH plane [21] or computation of additional semi-automated landmarks [22] have been shown to be reliable, but are more complex in terms of implementation.

Our third objective was to assess whether the conventional FH plane and the “new” FH plane were parallel. Our results regarding the vertical positions of IAF-L and IAF-R to the conventional FH plane, as well as the angular differences between the 2 planes, show that the planes are not parallel for most subjects. The vertical distance of IAF-L and IAF-R to the conventional FH plane showed significant variations in our cohort (Figure 3). This tends to invalidate the relevance of this “new” FH plane as a replacement for the conventional FH plane. Our results are not consistent with Pittayapat *et al*.’s findings, which reported an angular difference of 0.53 ± 0.37° between the conventional FH plane (called FH1 in their study and constructed using mid-Po, Or-R, Or-L) and their new FH plane. In order to facilitate the comparison between our data and Pittayapat *et al*.’s, the angular distances between the conventional and all newly proposed FH planes are provided in Supplementary Materials 3. The discrepancies between the two studies might be explained by a different definition of the IAF-R and IAF-L points. We tried our best to follow the instructions given by the aforementioned authors to define these points (Supplementary Materials 1), but a more precise anatomical description might be needed. The average duration of CT scan landmarking confirms that this is a time-consuming procedure, requiring prior training of the operators and potentially limiting clinical implementation. Our durations were in line with Hassan *et al*.’s results, who reported an average time of 10:41 ± 4:01 minutes to localize 22 landmarks using a similar protocol [13]. Given the operator training and time needed to place the 3D landmarks manually, semi- or fully automatic landmarking could help advance clinical use of 3D cephalometry [31].

This study has three main limitations. Firstly, it was based on a retrospective selection of a limited number of clinical cases, which can be a source of potential bias or imprecision in the statistical analysis. Despite the heterogeneity of our sample (sex, age, skeletal classes), we assumed that the within-subject standard deviation of each landmark was the same for all the patients [32]. We believe that our randomly-selected cases from clinical practice are an asset for the clinical applicability of our results because they are representative of the variability encountered in a clinical practice. Secondly, it was performed on CT scans instead of CBCT scans, which are more commonly used for 3D cephalometric studies. We made this choice because CT scanning is the imaging modality currently used for orthognathic surgery planning in our department. We chose to include only presurgical patients in this study because they display a variety of significant craniofacial deformities for which 3D cephalometry could be very beneficial for in-depth evaluation [1,33]. Thirdly, the placement of most of our landmarks was carried out on the CT scans’ 3D surface models, using the MPR views for adjustments and verifications. As has been reported previously, while the use of 3D surface models make the annotation process easier and more robust, it implies prior segmentation of the CT scans [13]. Performing the segmentation process manually is tedious and time-consuming, but full automatization of the task, an active and promising research field, could resolve this problem [34]. Given that most of the annotations were made on 3D surface models, we hypothesize that using CBCT scans instead would yield similar results. In order to evaluate the consequences of our results on patients’ soft tissues, the 3D surface models used in this study could be superimposed with the patients’ facial scans [35]. The virtual patients obtained using this recently described technique could provide valuable additional clinical insights and help surgical planning. Not having a facial scanner at our disposal, we were not able to test the technique in our study.

Overall, our repeatability and reproducibility study on CT scans showed that “dental” and “foraminal” 3D landmarks tended to be more reliable than “conventional” cephalometric 3D landmarks in presurgical patients. Despite the poor overall reliability of the landmarks orbitale and porion in 3D, the conventional FH plane is a reliable horizontal reference for head reorientation and vertical measurements. The new FH plane, using IAF instead of porion, provided a reliable horizontal reference but was not found to be parallel to the conventional FH plane.

## Data Availability

The data underlying this article will be shared on reasonable request to the corresponding author.

## Supplementary Materials

- Supplementary Materials 1: Written instructions for the landmarking process on the Mimics software.
- Supplementary Materials 2: Analysis of outliers.
- Supplementary Materials 3: Angular distances between conventional and novel FH planes.

## Acknowledgment

The authors would like to thank the “Association Les Chirurgiens Maxillo-Faciaux” for its technical support.

## Data availability

The data underlying this article will be shared on reasonable request to the corresponding author.

## Ethical Approval

The IRB “Comité d’Ethique pour la Recherche en Imagerie Médicale” (CERIM) gave ethical approval for this research (number CRM-2001-051).

