## Supplementary Materials for "Three-dimensional cephalometric landmarking and Frankfort horizontal plane construction: reproducibility of conventional and novel landmarks"

Supplementary Material 1

#### Written instructions for landmarking process
#### Mimics software

Useful Mimics commands:

| | | |
|---|---|---|
| 3D view | Rotation of the object | Mouse right click + move near the object |
| | Tilt of the object | Mouse right click + move far from the object |
| | Translation of the object | Shift + right mouse click + move |
| | Zoom in/out | Ctrl + right mouse click + move<br>or Mouse scroll |
| 2D views | Translation of the slice | Shift + right mouse click + move |
| | Zoom in/out | Ctrl + right mouse click + move |
| | Change slice | Mouse scroll |
| | Change contrast | Alt + right mouse click + move |

### Procedure

- Open Mimics document
- If needed, change document’s layout to get a bigger 3D view: View > Layouts > Horizontal
- On the right panel, hide all masks (eye icon) & all objects other than Upper Skull / Mandibula / Upper Teeth / Lower Teeth
- Orient the skull object for it to be approximately aligned with Frankfort plane
- Launch Analyze > Measure and Analyze and choose “Full ceph analysis”

- Start Time measurement (chronometer)

- Follow the order of the landmarks to annotate (1:, 2:, 3:.... to 33:)
- If a landmark is missing (e.g. missing tooth), do not annotate it

| | | |
|---|---|---|
| 1: Nasion | Medial (and upper) point of the fronto-nasal suture | 3D view, frontal<br>Check on 2D slices if necessary (axial/sagittal) |
| 2,3: Orbitale L/R | Lowest point of the orbitale rim L/R | 3D view, frontal and check from above that the points are well on the orbital rim |

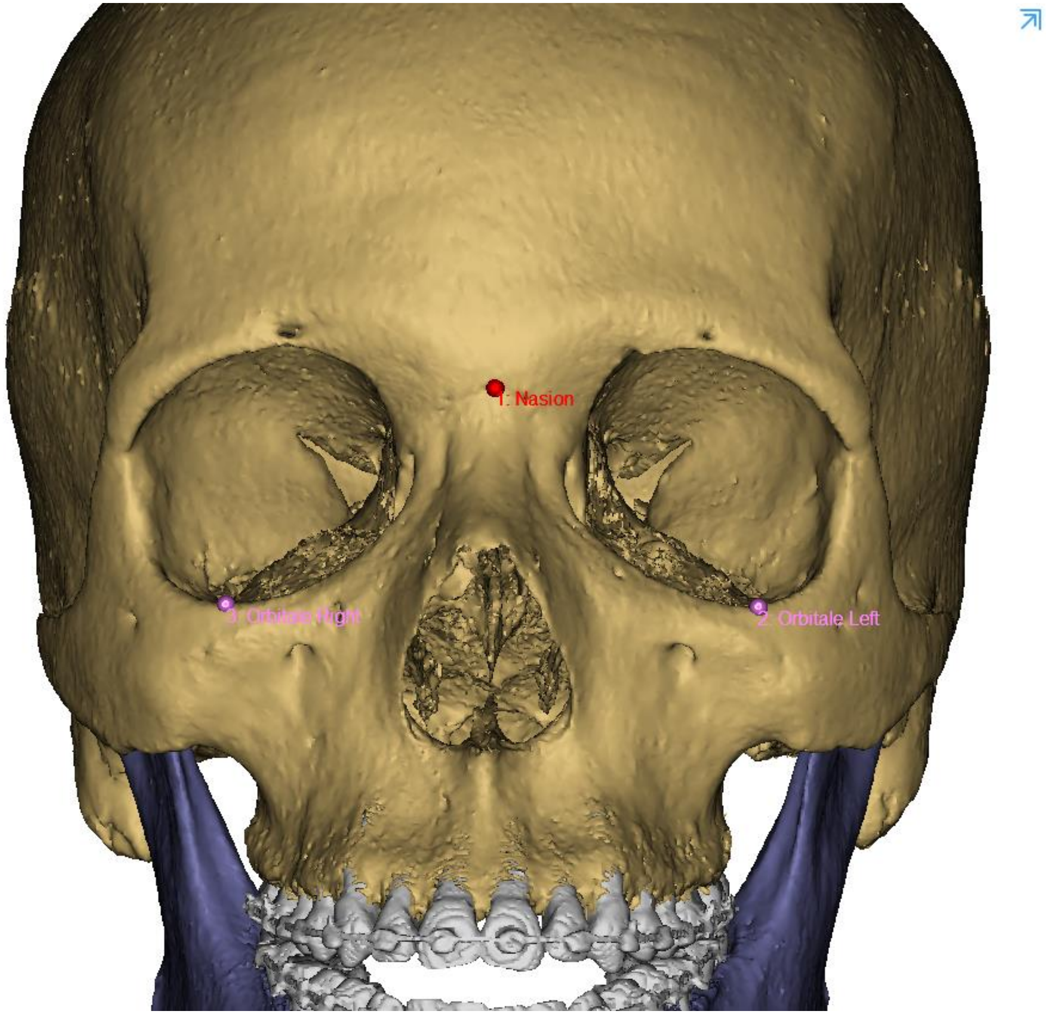

| | | |
|---|---|---|
| 4,5: Infraorbital Foramen L/R | External & most distal point of the infraorbital foramen L/R | 3D view, ¾ mesio-lateral view<br>Check on 2D slice (Axial) |

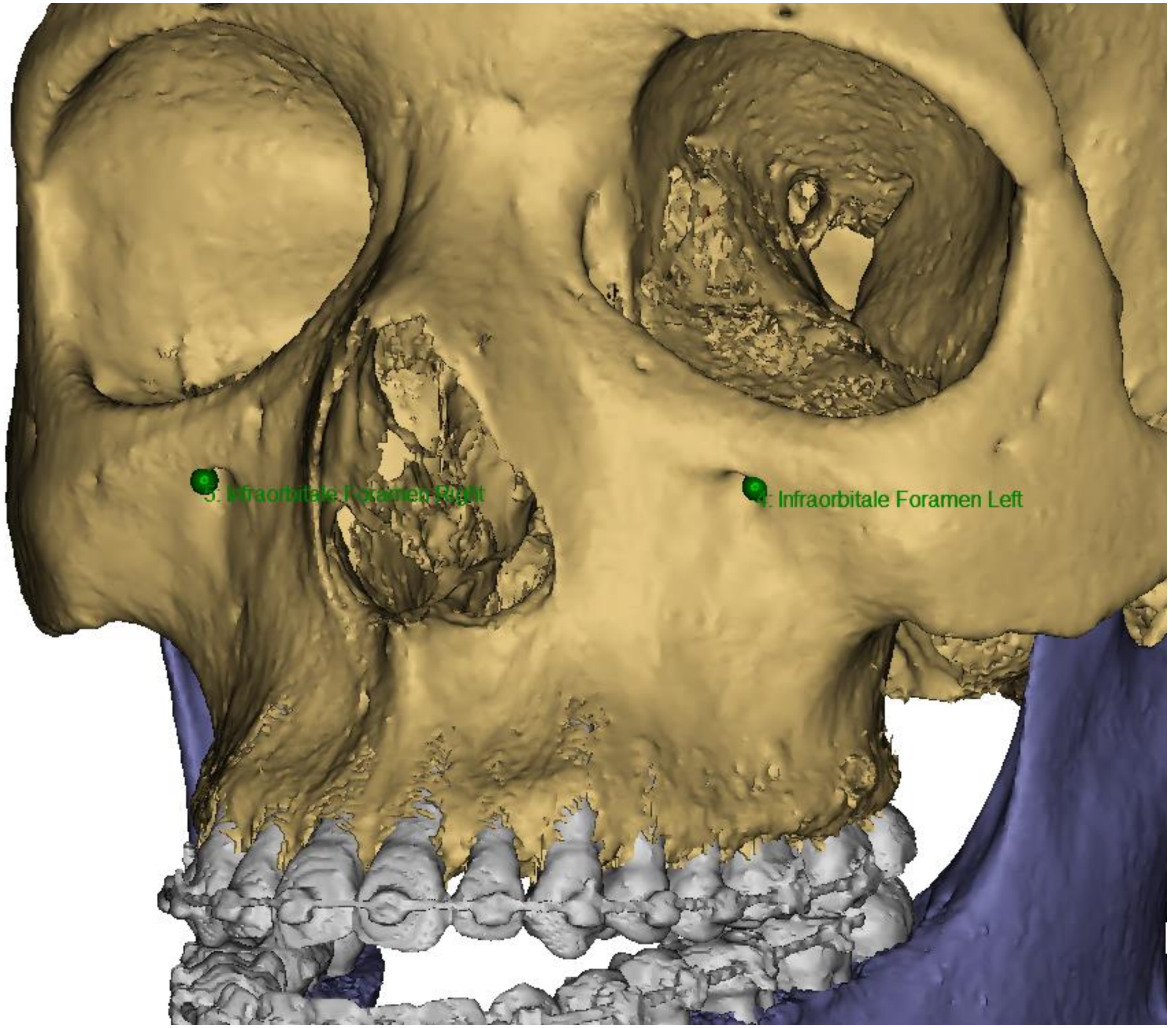


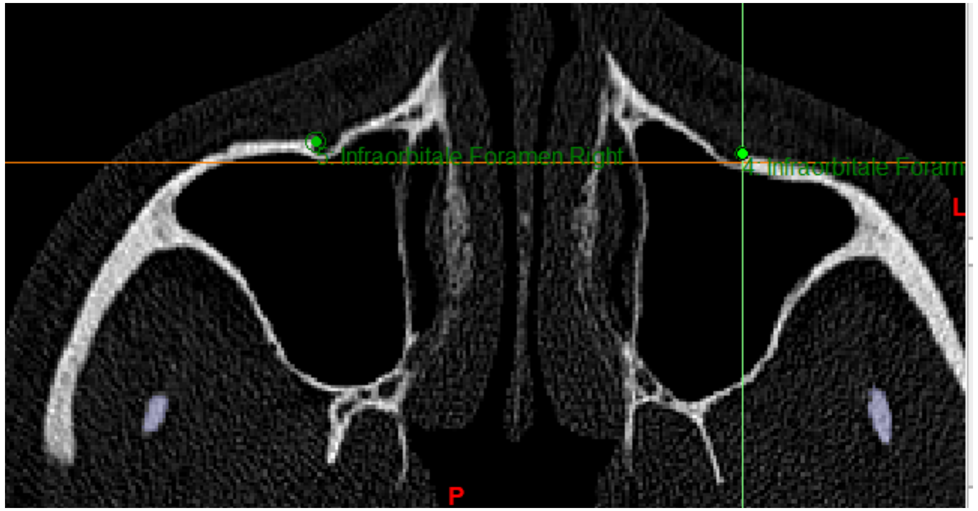


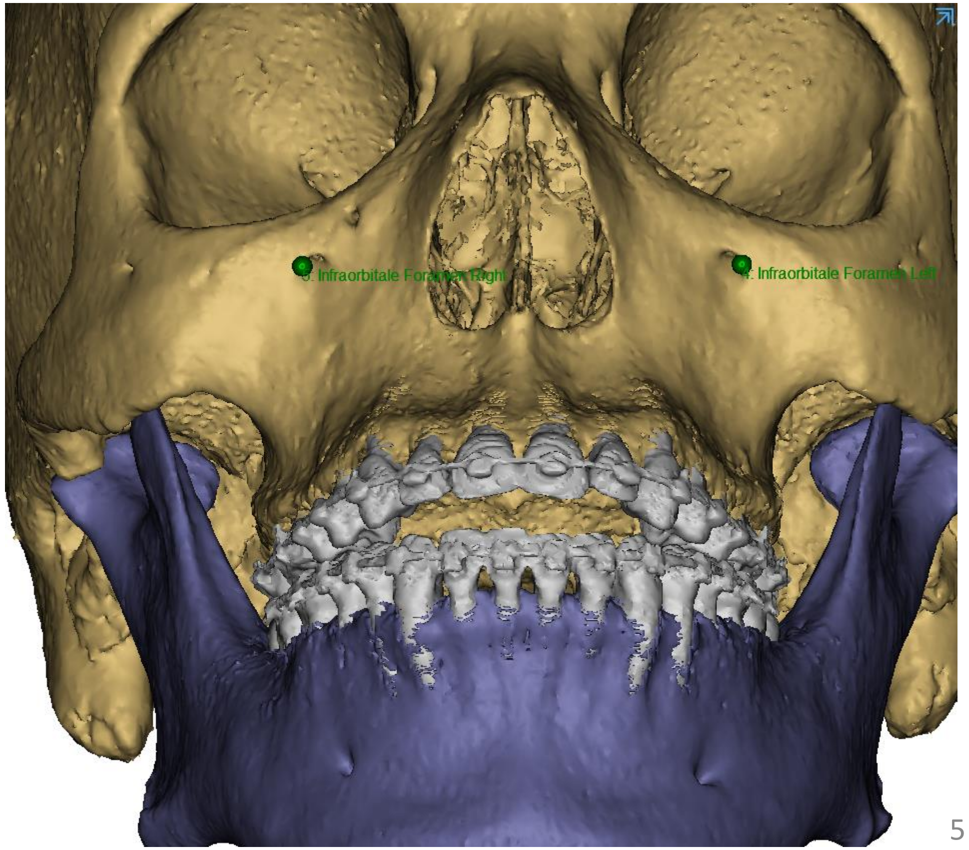

| | | |
|---|---|---|
| 6: ANS | Medial and most anterior point of the nasal spine | 3D view, frontal<br>Check on 2D slices (axial/sagittal)<br>In case of bifid spine, stay at a medial position |
| 7: A | Medial & most posterior point of the maxilla | 3D view, frontal<br>Check afterwards laterally (see after point 18) |
| 8: B | Medial & most osterior point of the mandible | 3D view, frontal<br>Check afterwards laterally (see after point 18) |
| 9: Pog | Medial and most anterior point of the mandible | 3D view, frontal<br>Check afterwards laterally (see after point 18) |
| 10: Gnathion | Medial & mid-point between Pog and Me | 3D view, frontal<br>Check afterwards laterally (see after point 18) |
| 11: Menton | Medial and lowest point of the mandible | 3D view, inferior<br>Check afterwards laterally (see after point 18) |

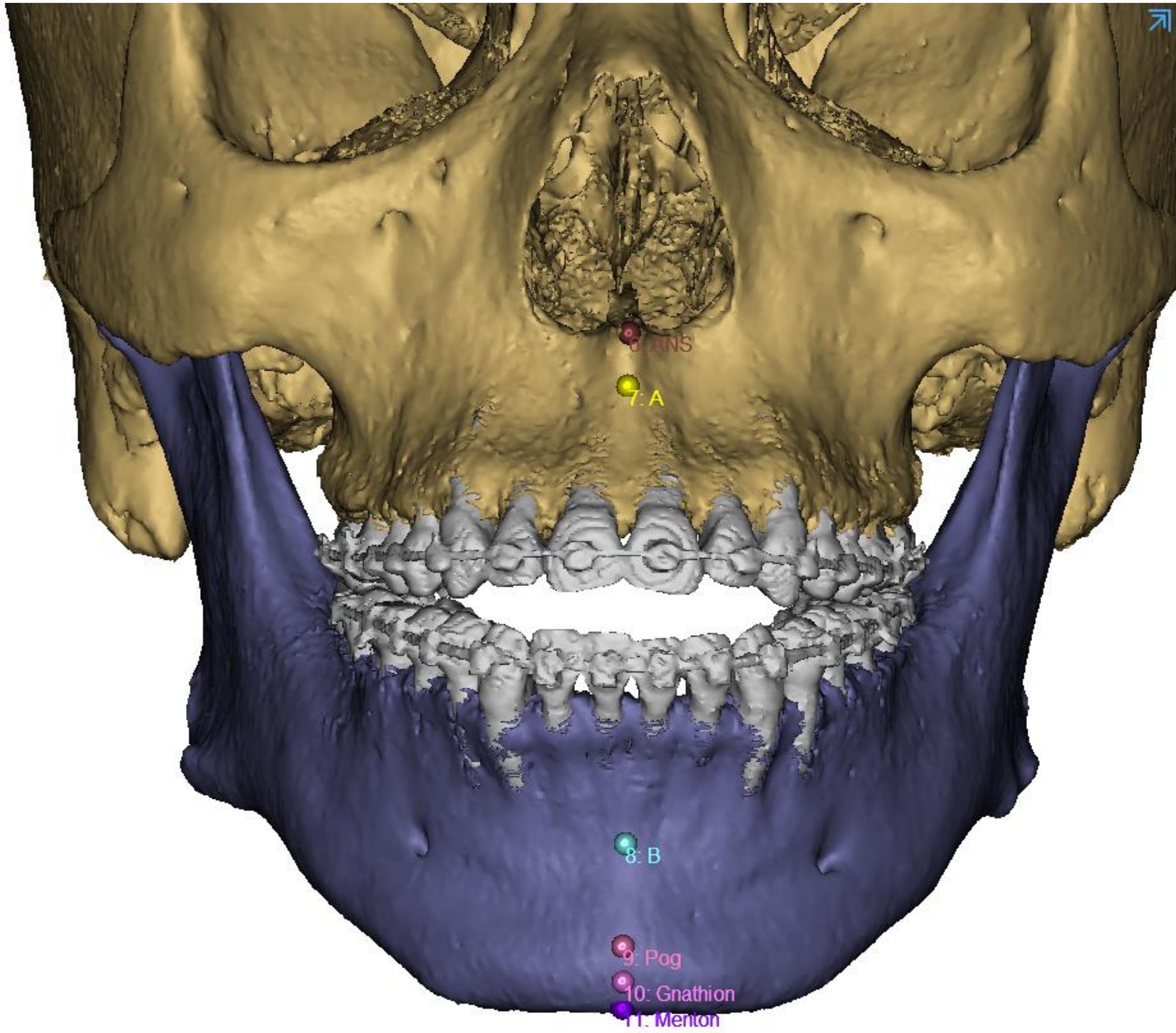

| | | |
|---|---|---|
| 12, 13: Mental Foramen L/R | External most mesial point of the mental foramen L/R | 3D view, ¾ disto-lateral view<br>Check on 2D slice (Axial) |

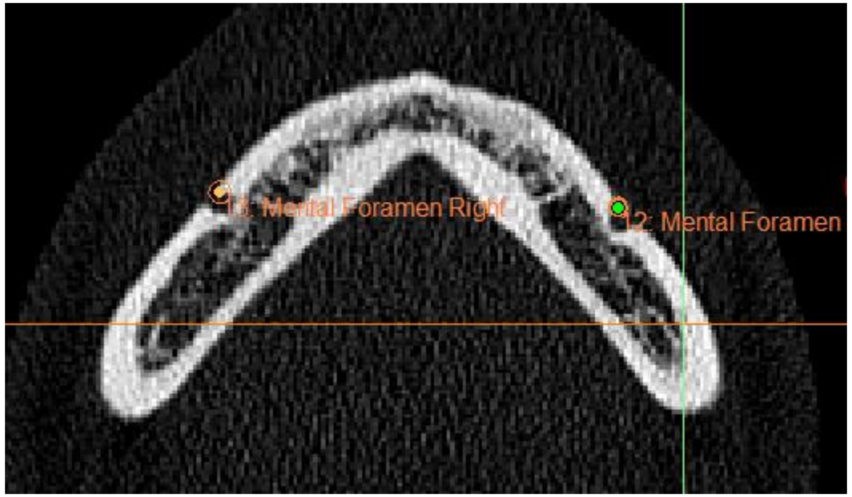


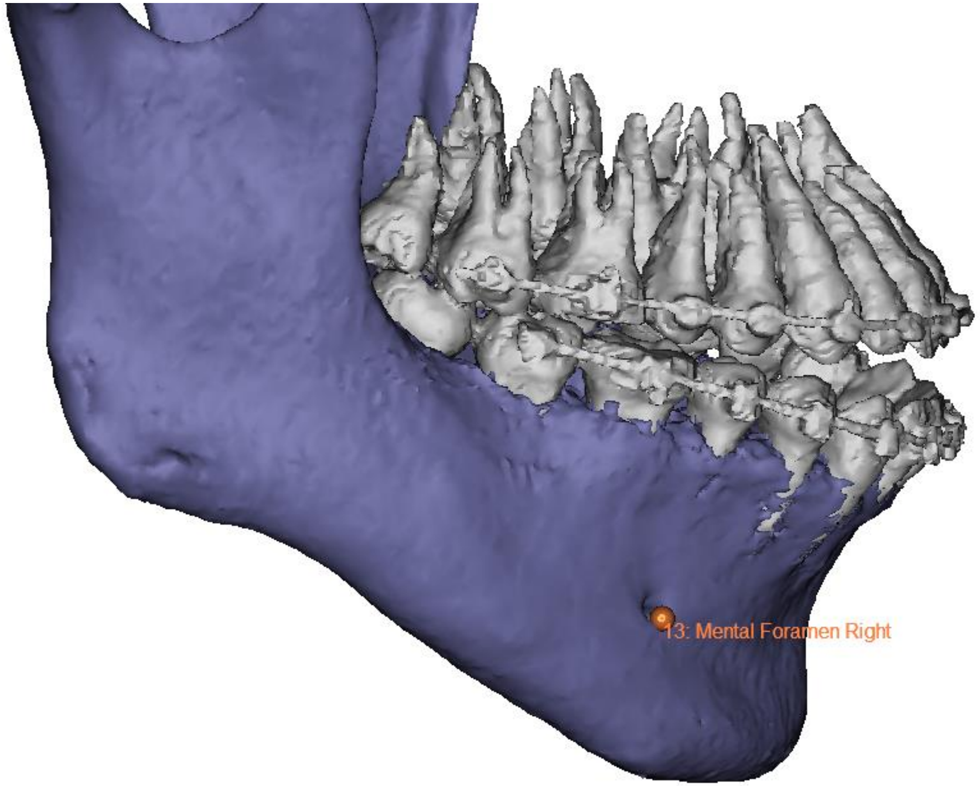


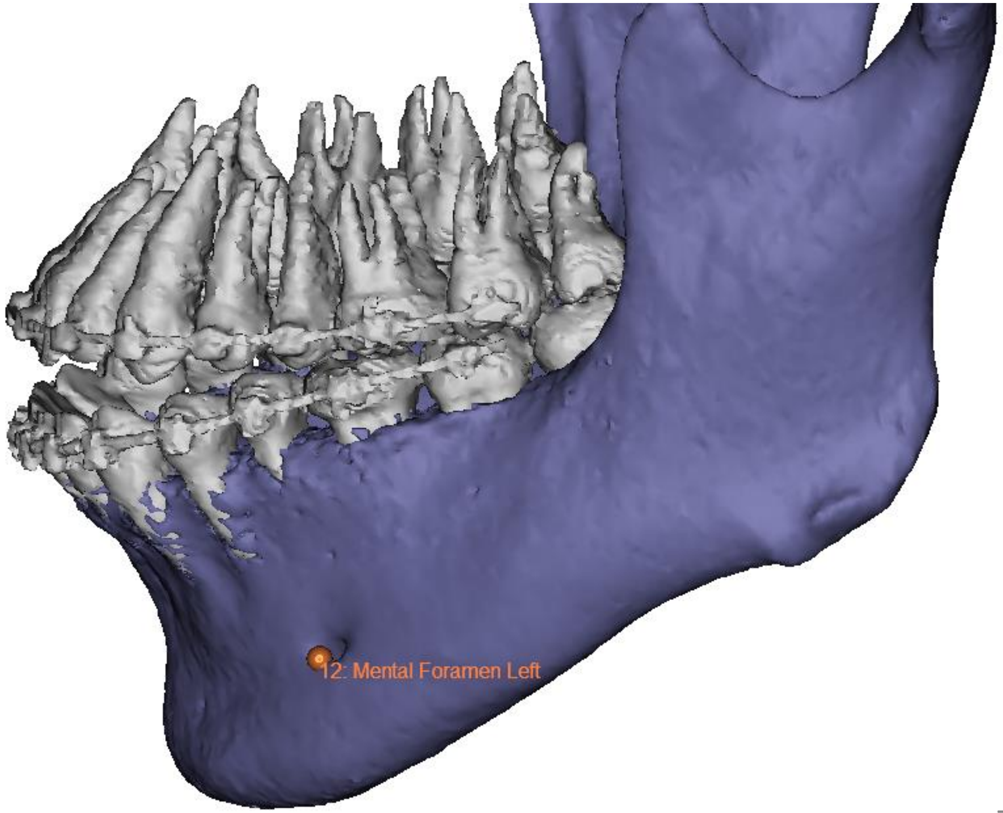

| | | |
|---|---|---|
| 14,15: Gonion L/R | Mid-point of the gonial angle L/R | 3D view, lateral. Mid-point between the beginning and the end of the mandibular angle<br>Check from behind that the points are well located on the mandibular external border<br>Before placing R one it can be easier to hide L one |

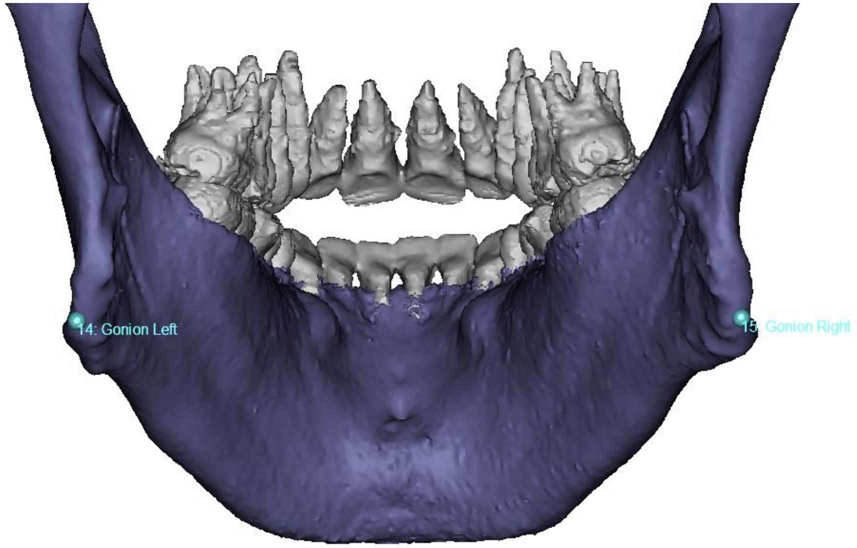


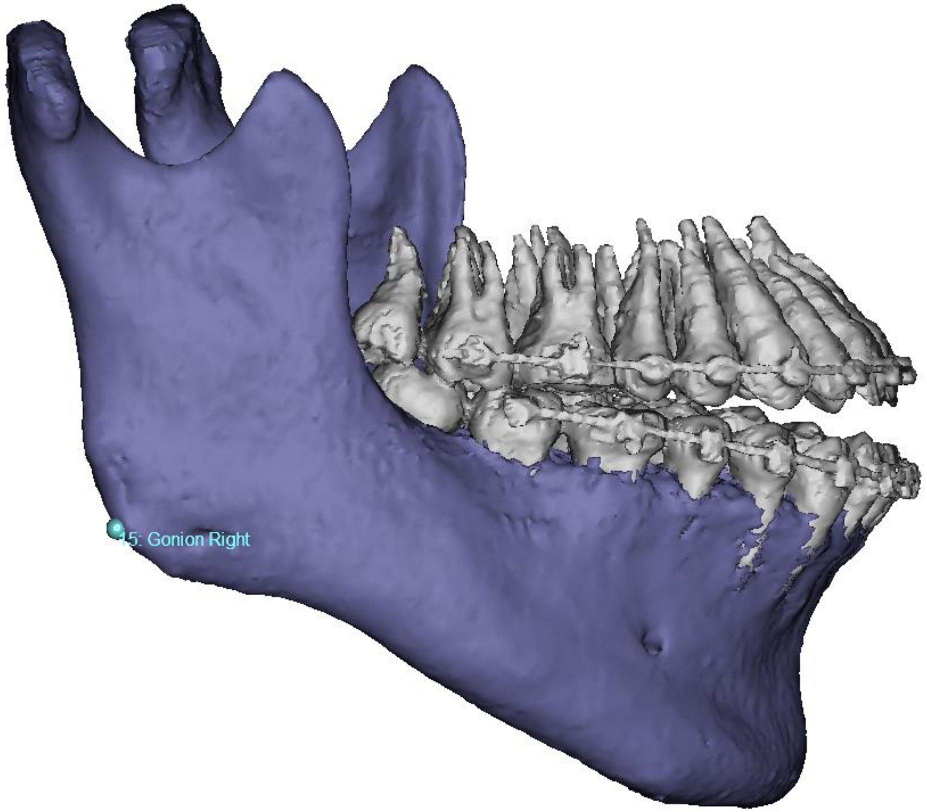


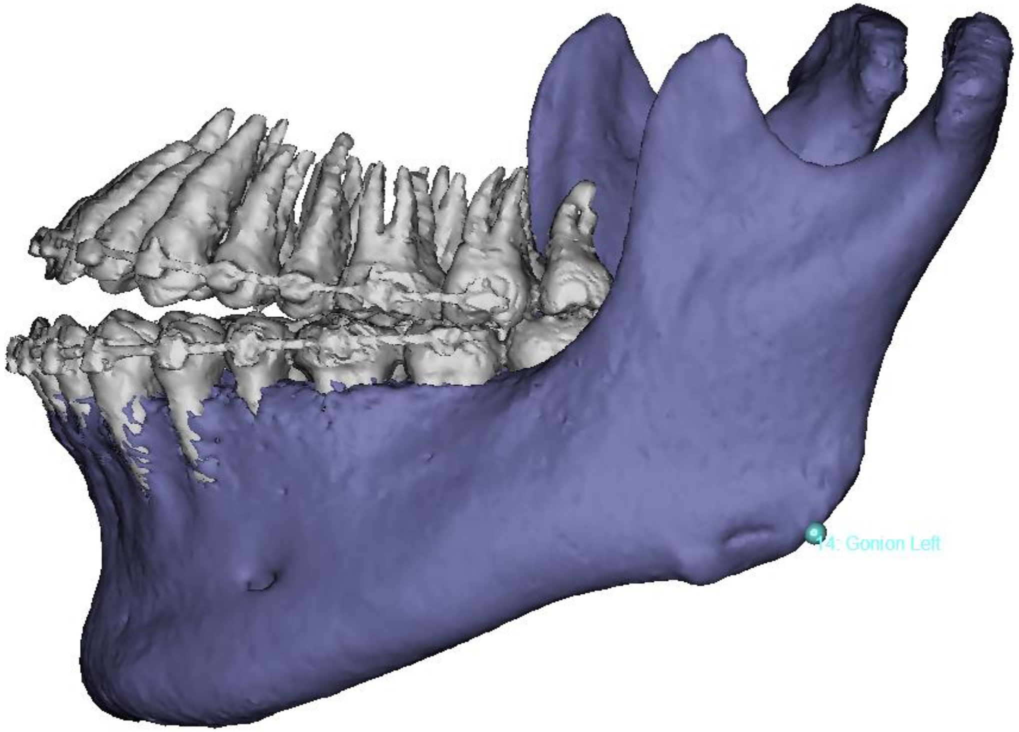

| | | |
|---|---|---|
| 16, 17: Porion L/R | External & uppermost point of the auditory canal L/R | 3D view, inferior. At the entry of auditory canal |

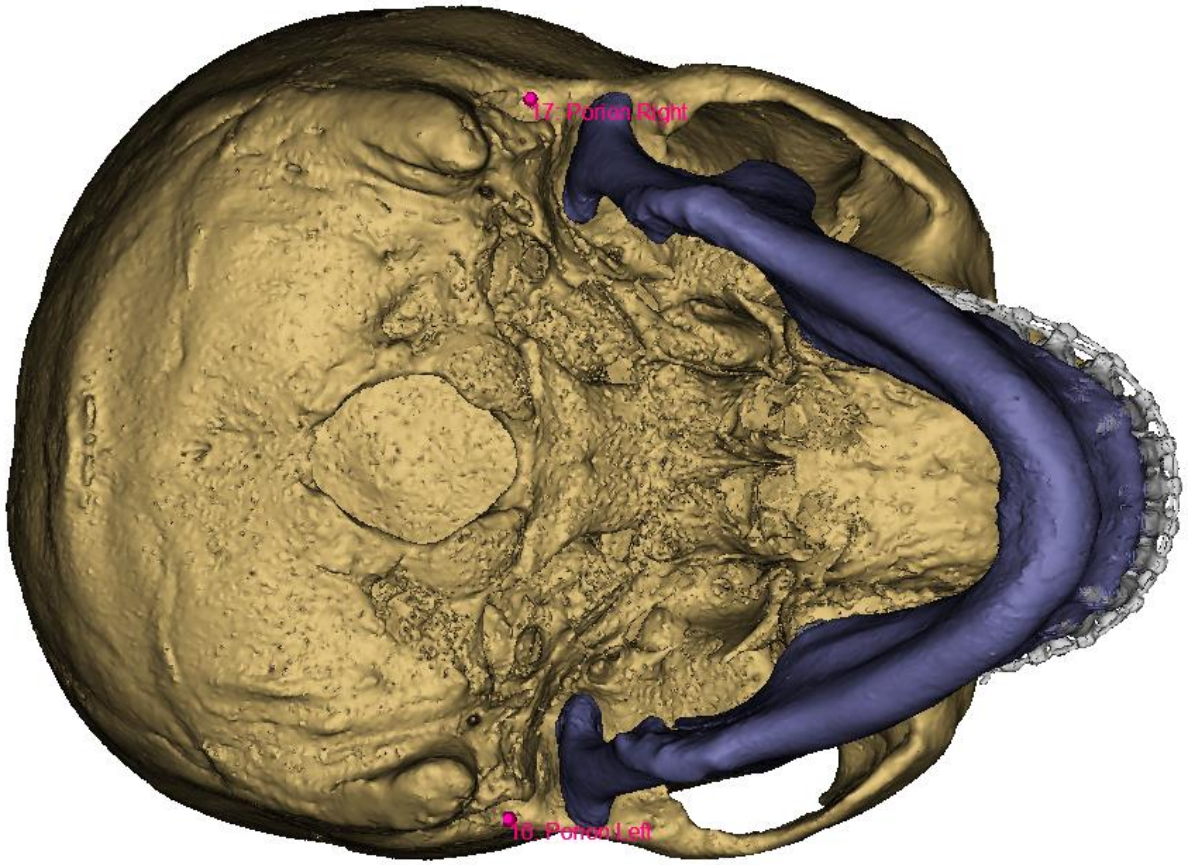


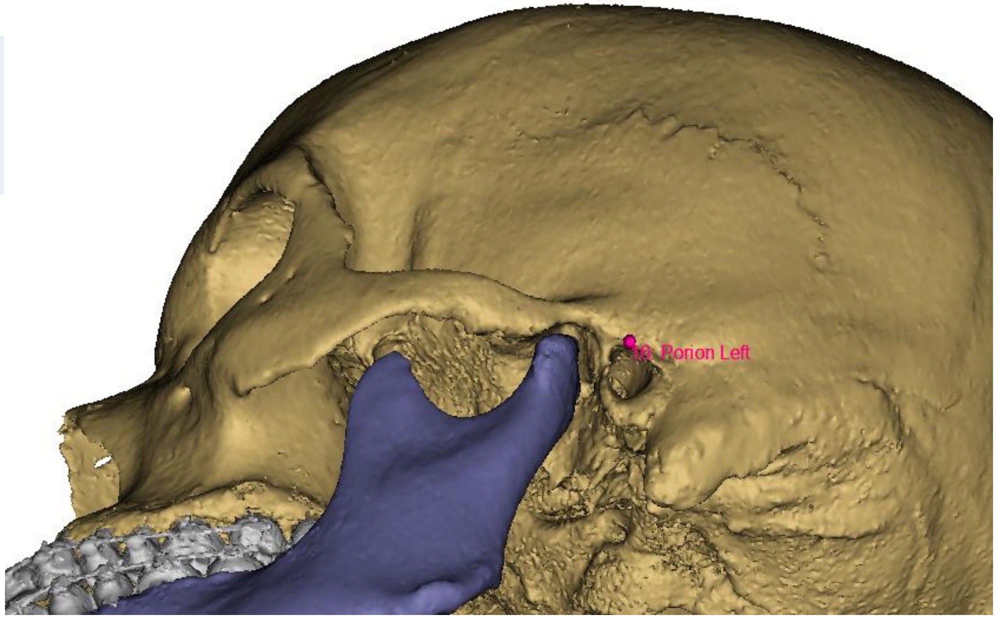


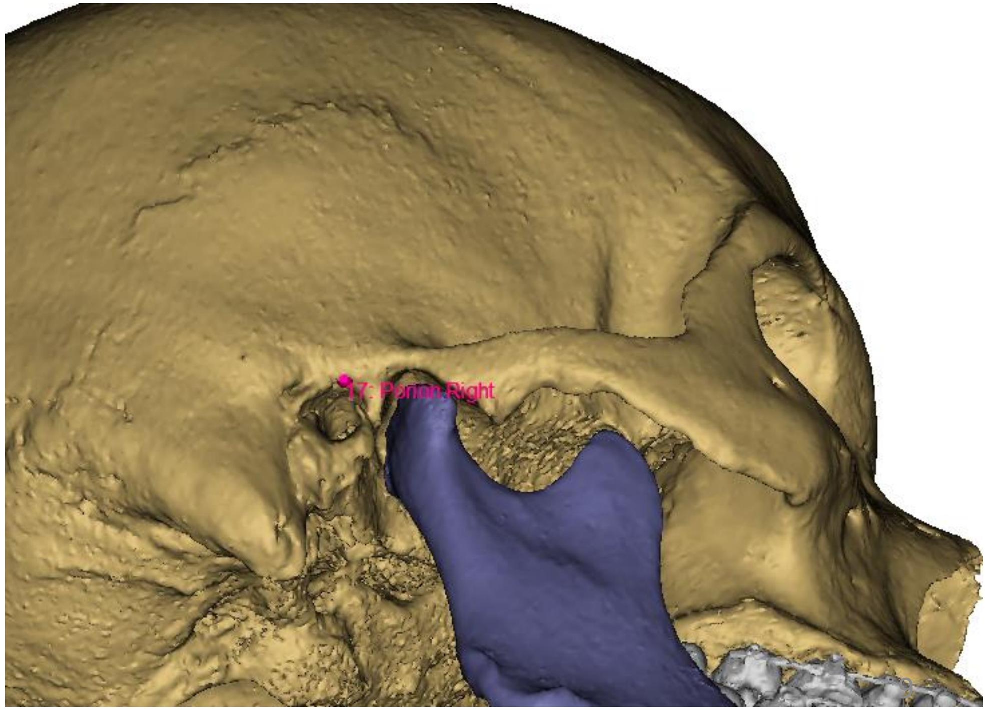

| | | |
|---|---|---|
| 18: PNS | Medial & most distal point of the osseous palate | 3D view, inferior-posterior<br>Check on 2D slice (sagittal) |

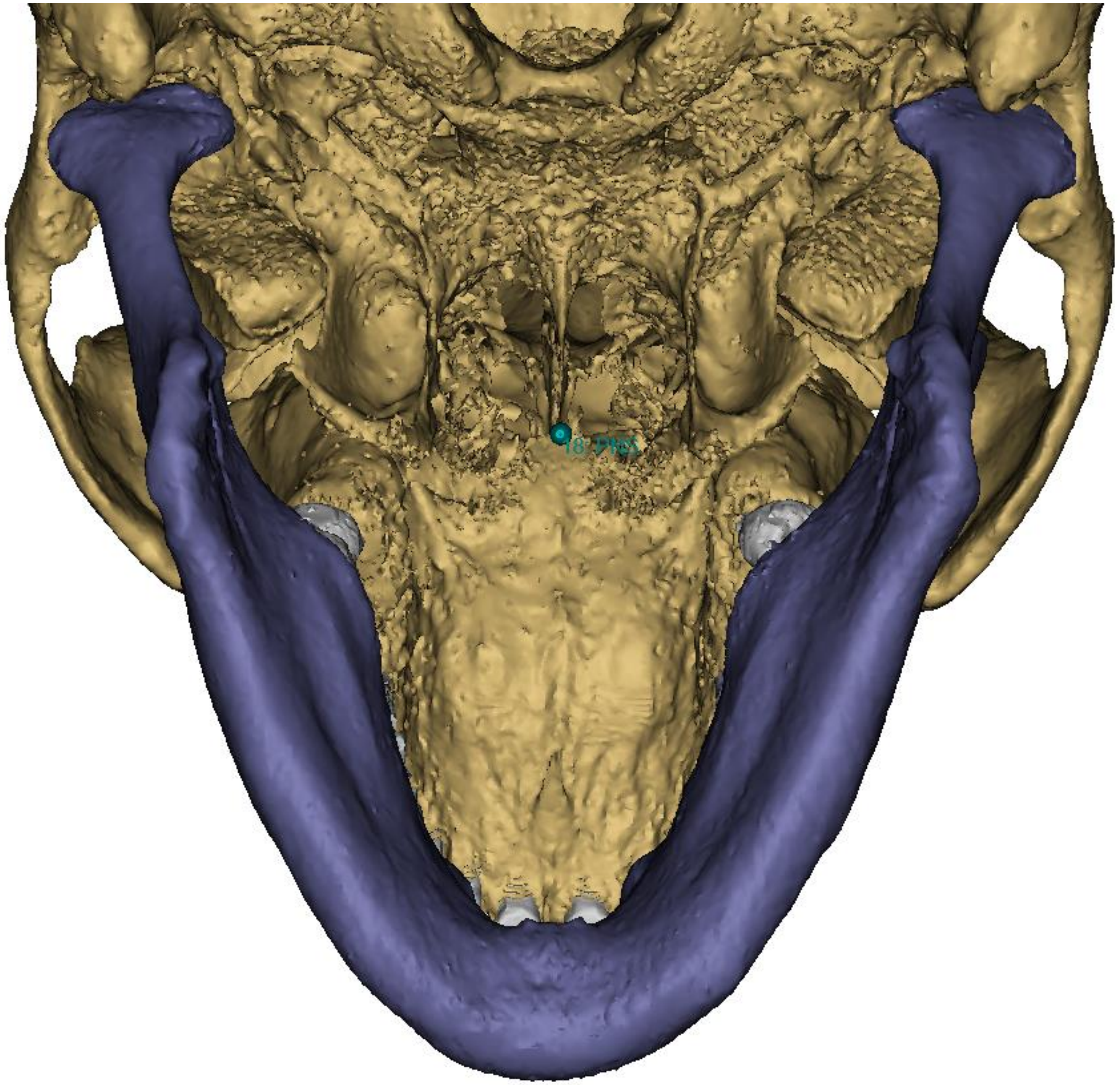


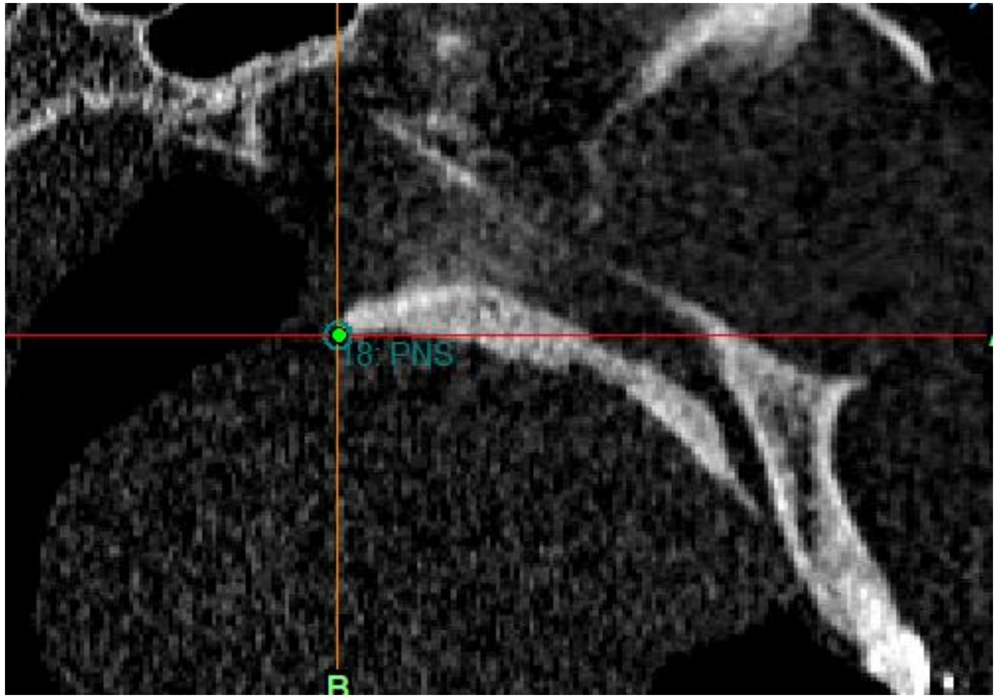

In the right lateral panel, hide the skull and align the 3D lateral view on the Frankfort plane automatically shown in red. **Check the landmarks 7, 8, 9, 10, 11 laterally:**

1/ 3D view, lateral, reorient to have Frankfort plane horizontal

For A point, show the skull

Check position of the points, correct them if necessary.

This can be done on the 2D slices (sagittal)

2/ 3D view, frontal, check that the medial position and alignment of the points is still OK

Afterwards, hide Frankfort plane (glasses icon) and show the skull

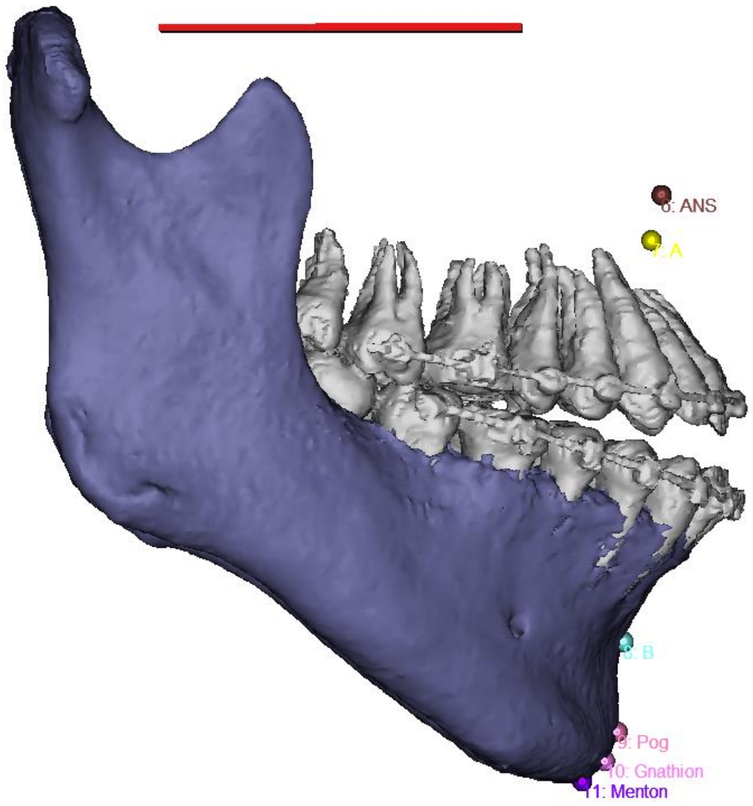


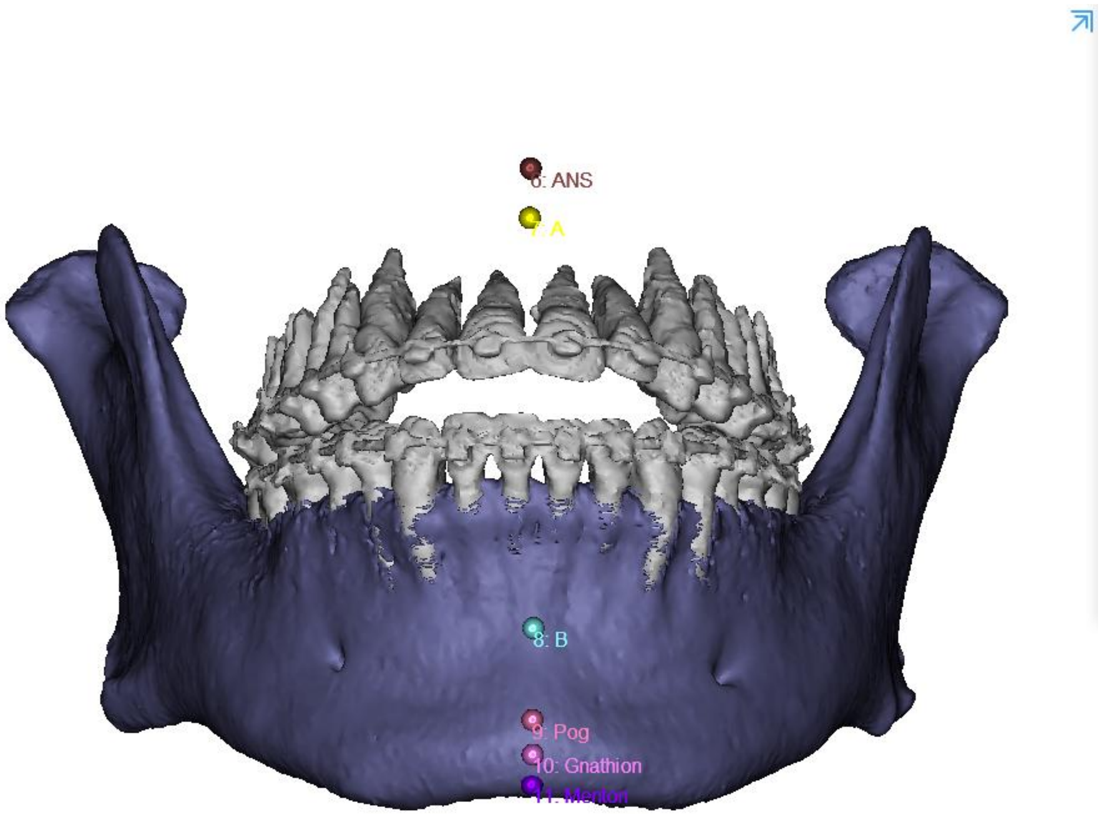

| | | |
|---|---|---|
| 19, 20: Internal Acoustic Foramen L/R | External, most mesial and posterior point of the acoustic foramen L/R | 3D view, ¾ postero-mesio-lateral view<br>Check on 2D slice (Axial) |

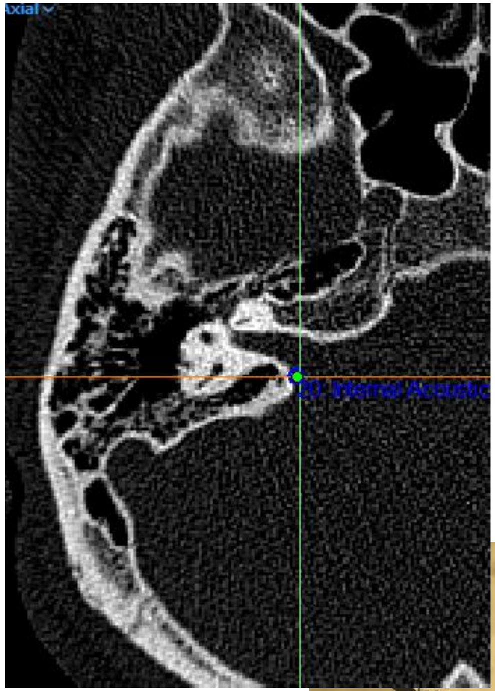


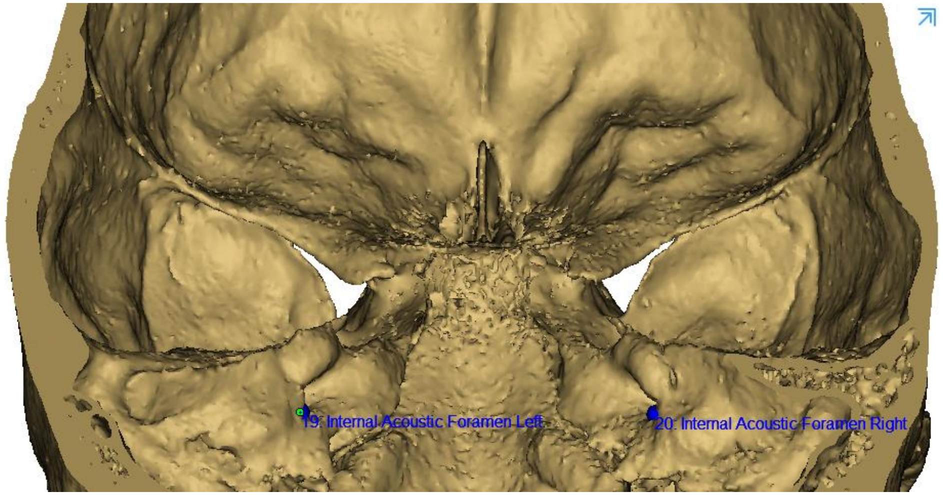


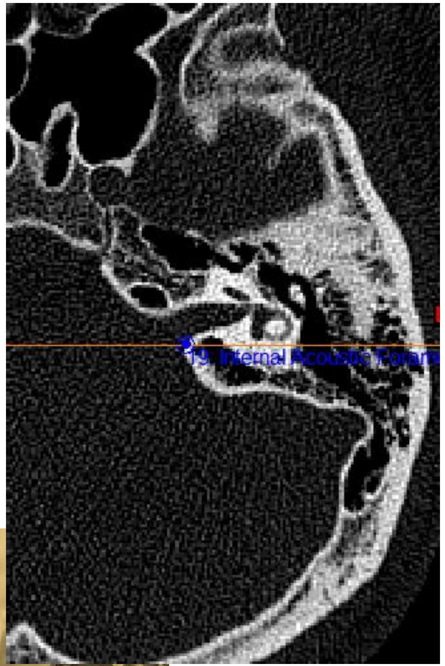


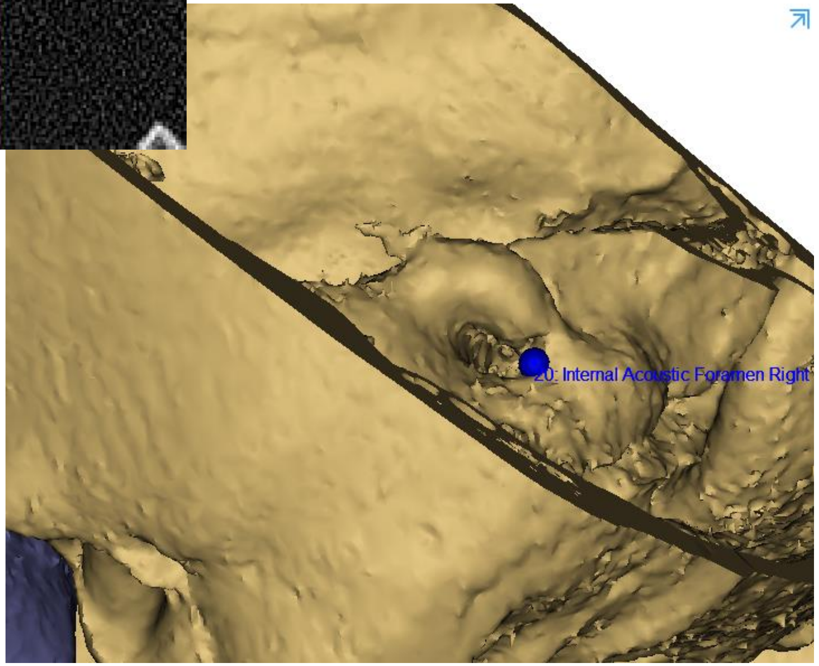


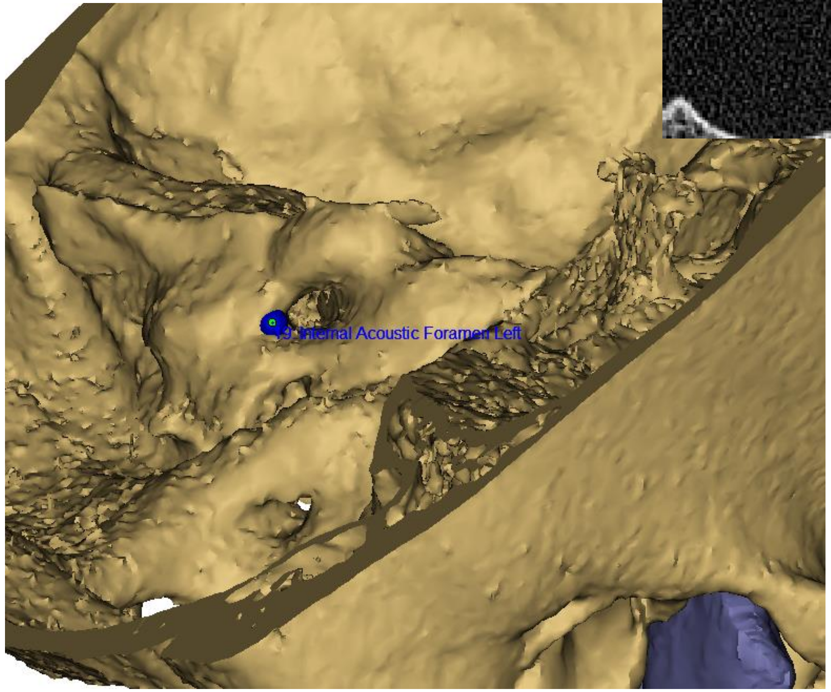

| | | |
|---|---|---|
| 21: Sella | Central point of the sella | 2D slices, first sagittal and refine on axial & coronal slices |

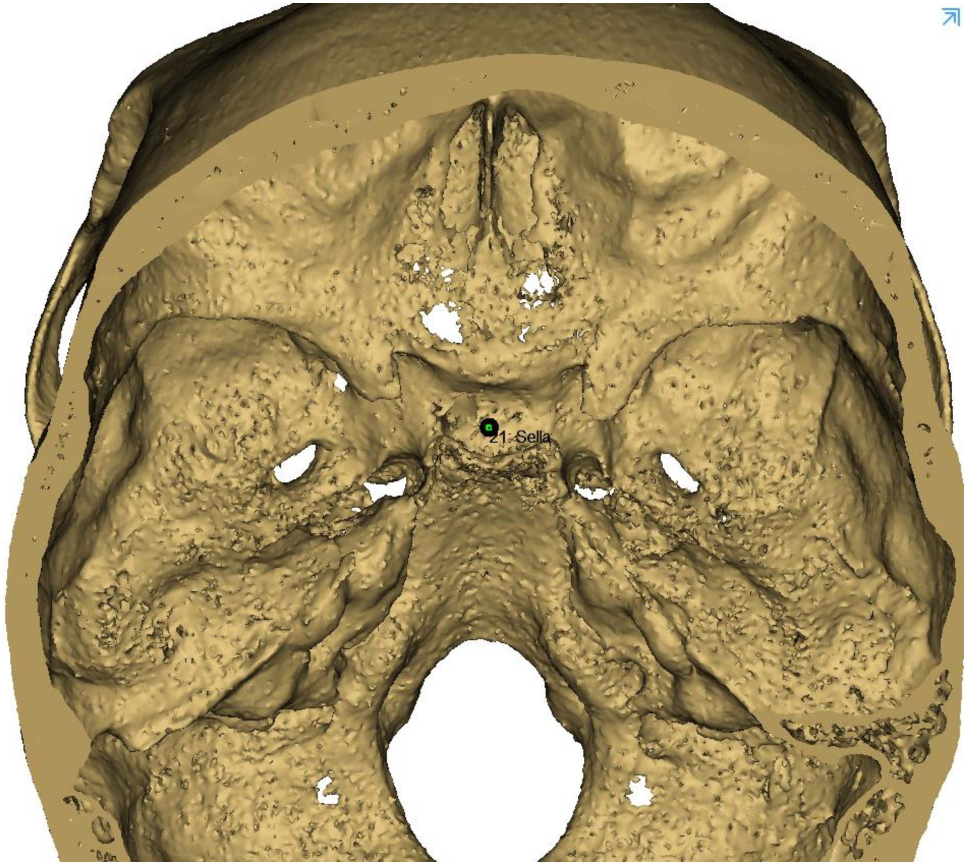


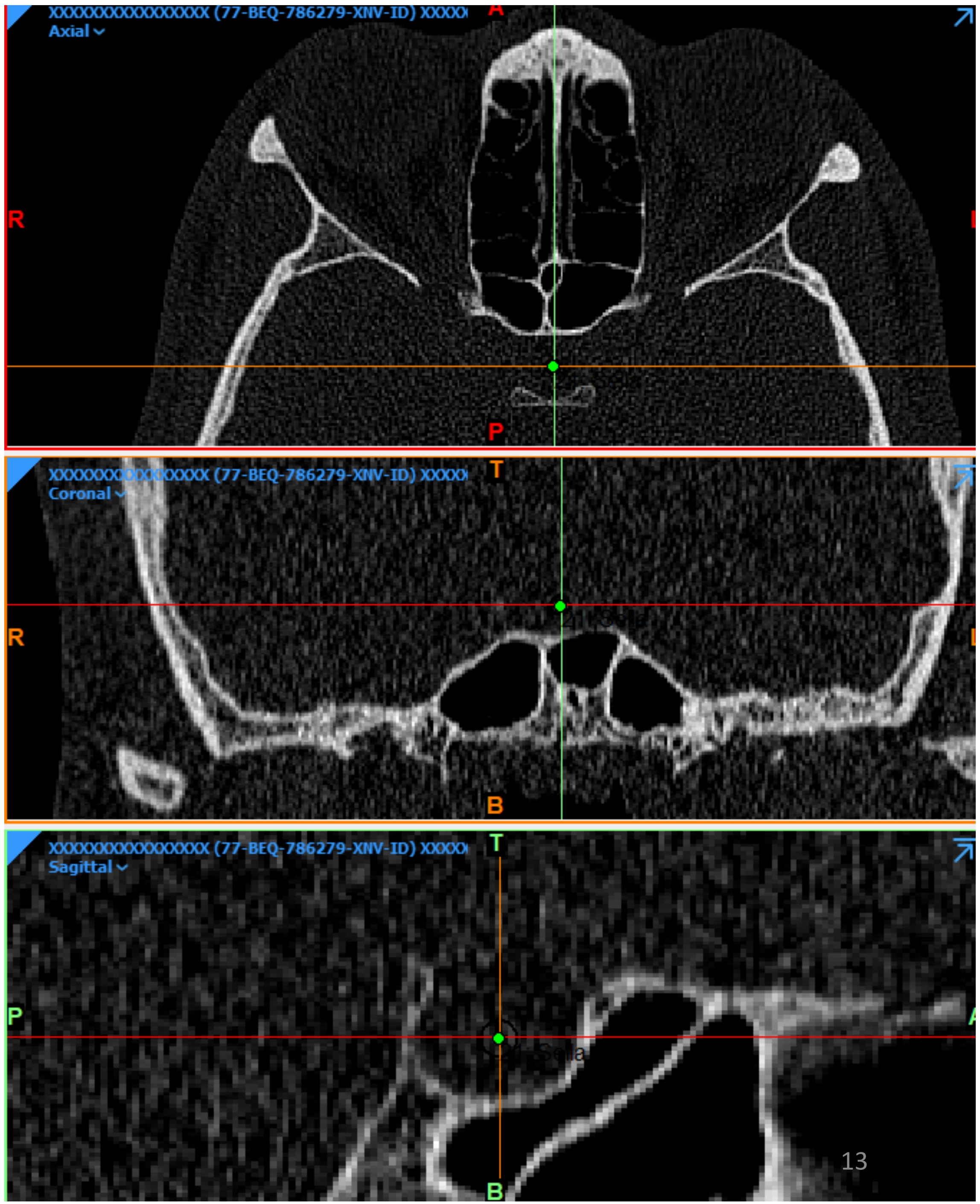

Hide all the landmarks localized (glasses icon)
Hide Upper Skull, Mandible and Lower teeth objects

| | | |
|---|---|---|
| 22, 23: 11E, 21E | Mid-point of 11/21 incisal edges | 3D view, inferior |
| 24,25: 16O, 26O | Summit of the mesio-palatal cusp 16/26 | 3D view, inferior |

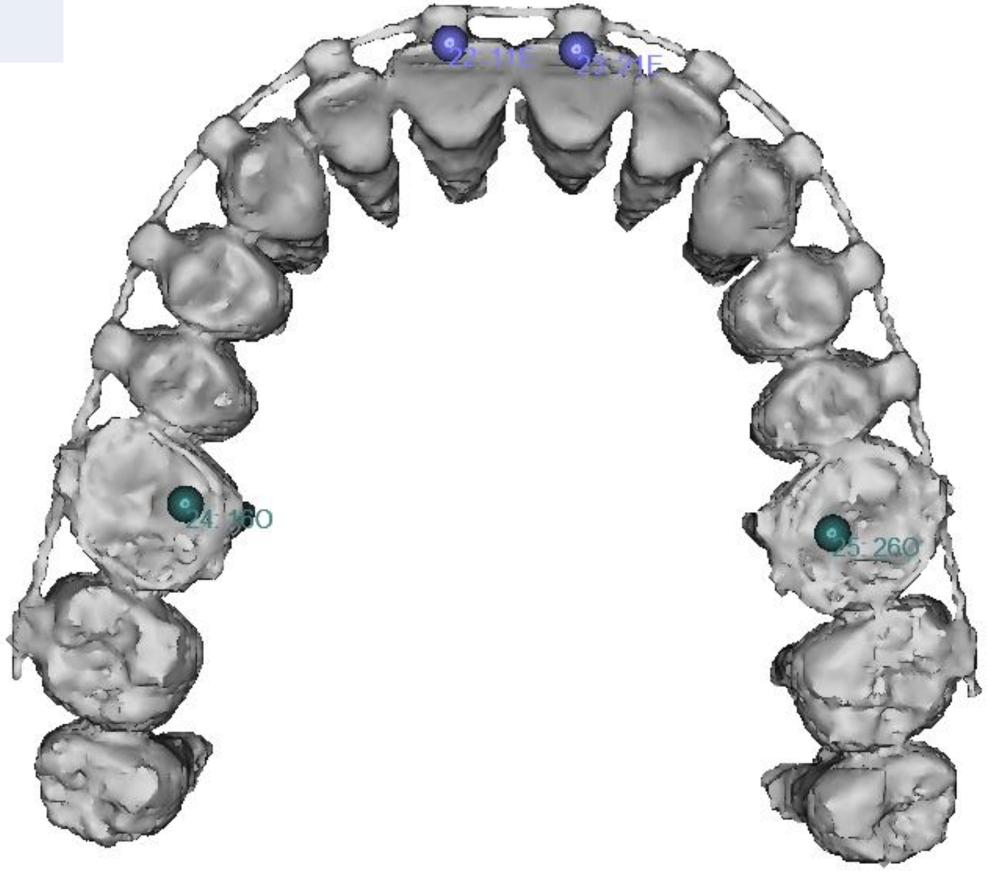

| | | |
|---|---|---|
| 26, 27: 11A, 21A | Root apex of 11/21 | 3D view, superior |

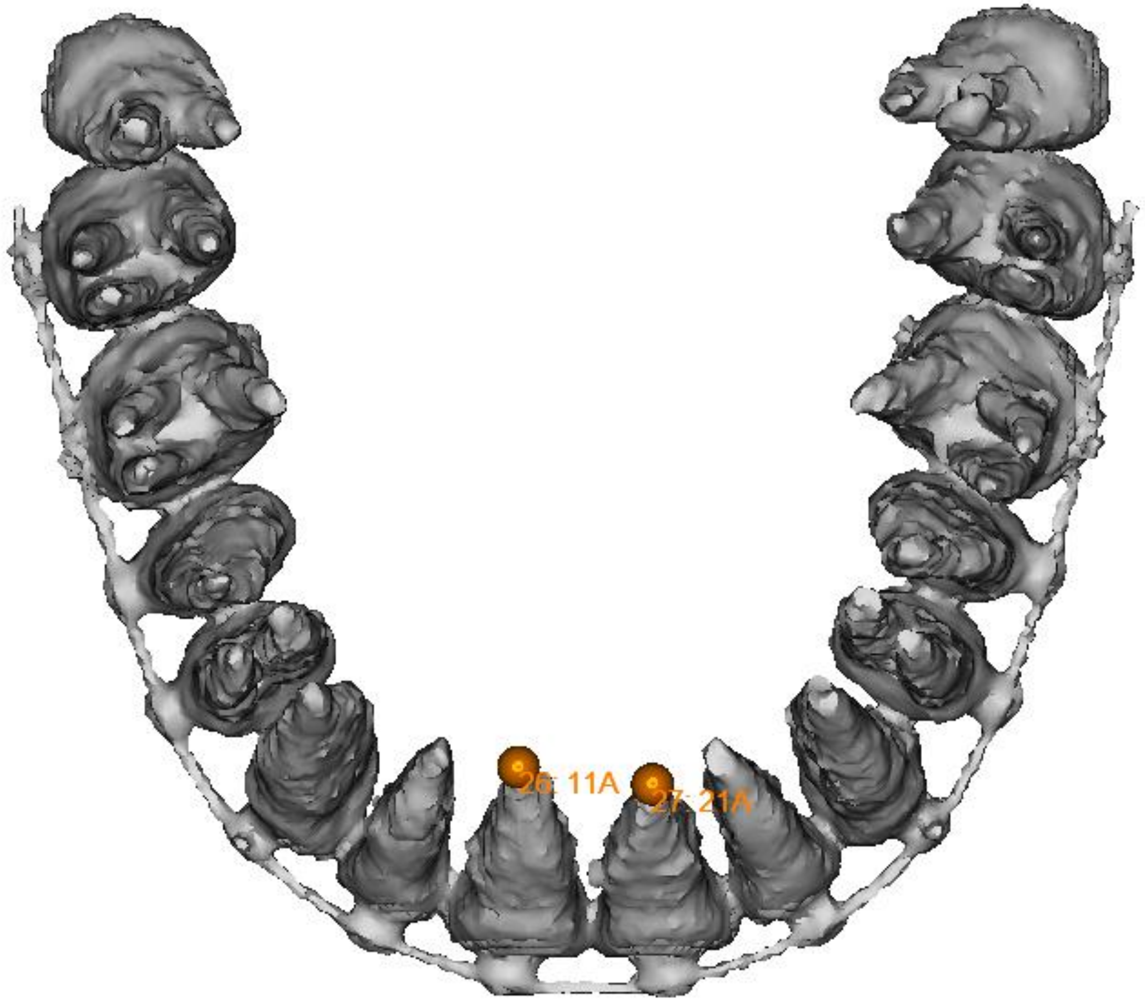

| | | |
|---|---|---|
| 28, 29: 31E, 41E | Mid-point of 31/41 incisal edgess | 3D view, superior |
| 30, 31: 360, 460 | Central fossa of 36/46 | 3D view, superior |

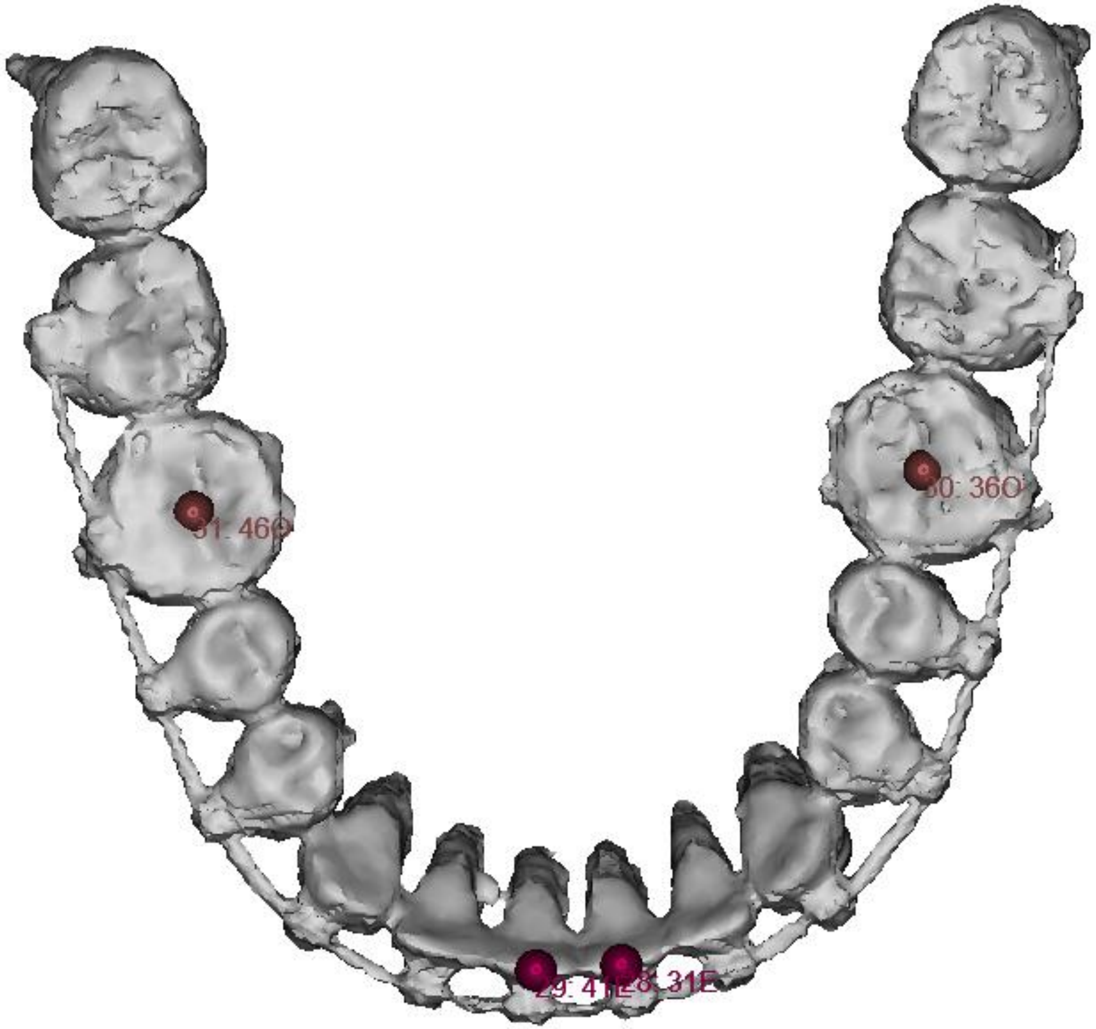

| | | |
|---|---|---|
| 32, 33: 31A, 41A | Root apex of 31/41 | 3D view, inferior |

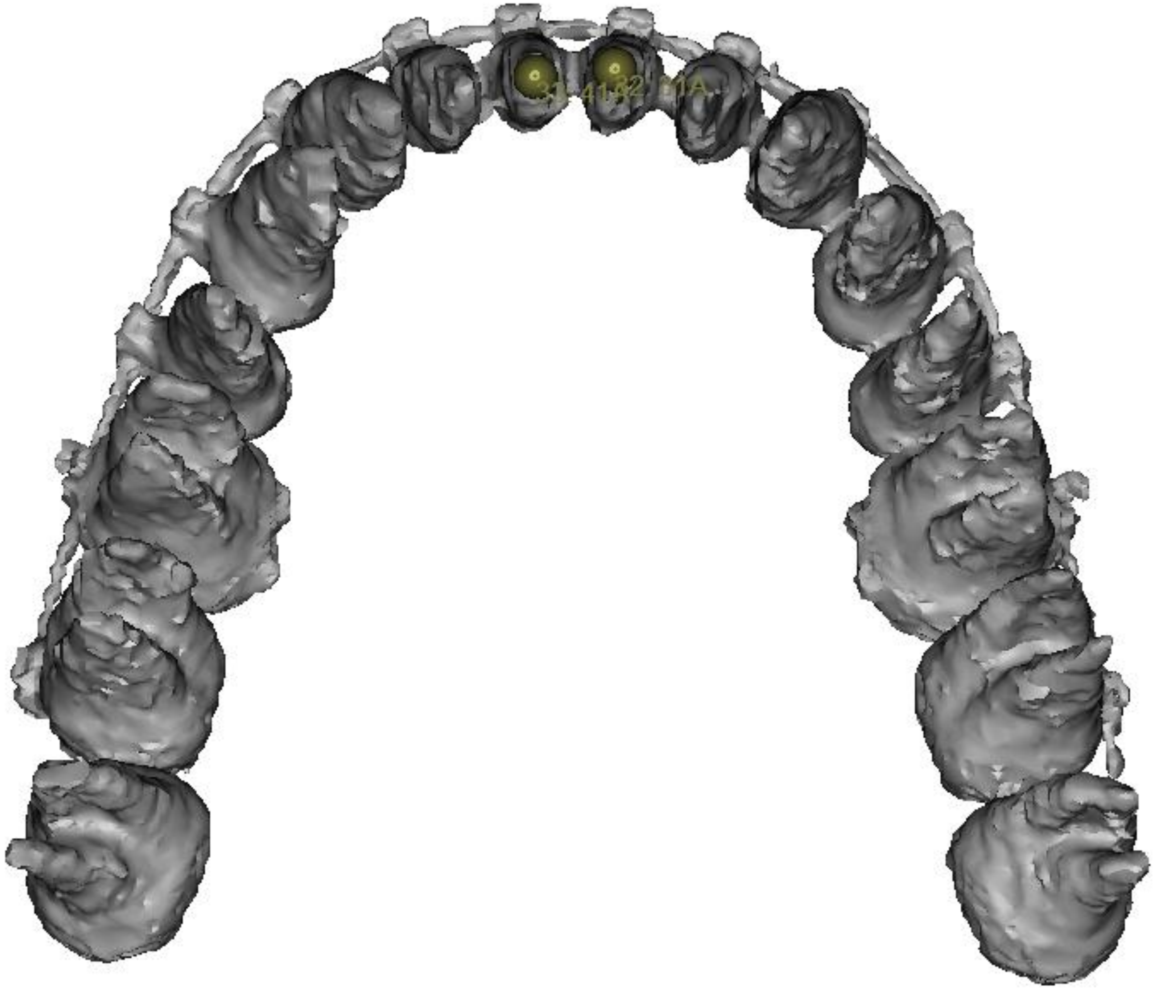

- Stop Time measurement (chronometer)

- Export the xml file using the “export” option of Measure & Analyze window

**Thank you ☺**

**Reproducibility of three-dimensional landmarking and Frankfort horizontal plane construction: a comparison of conventional and novel landmarks**

Supplementary Materials

- Supplementary Materials 2: Analysis of Outliers
- Supplementary Materials 3: Angular distances between conventional and novel FH planes

**Supplementary Materials 2**: Analysis of outliers

**Method.** For each landmark, repeatability and reproducibility standard deviations (SD) were computed according to the ISO 5725 standard (24) of the International Organization for Standardization. Upon first inspection of the results, the standard's recommendations were followed for clear outlier points, whose annotations were considered as missing data.

**Results.** We inspected all modified Bland-Altman graphs in order to find possible clear outliers. The only outliers were found for mental foramen points right/left localized by operator #3 during the first annotation session (subjects 4 to 20). Figure SM1 shows Bland-Altmann graphs of the results with these outliers.

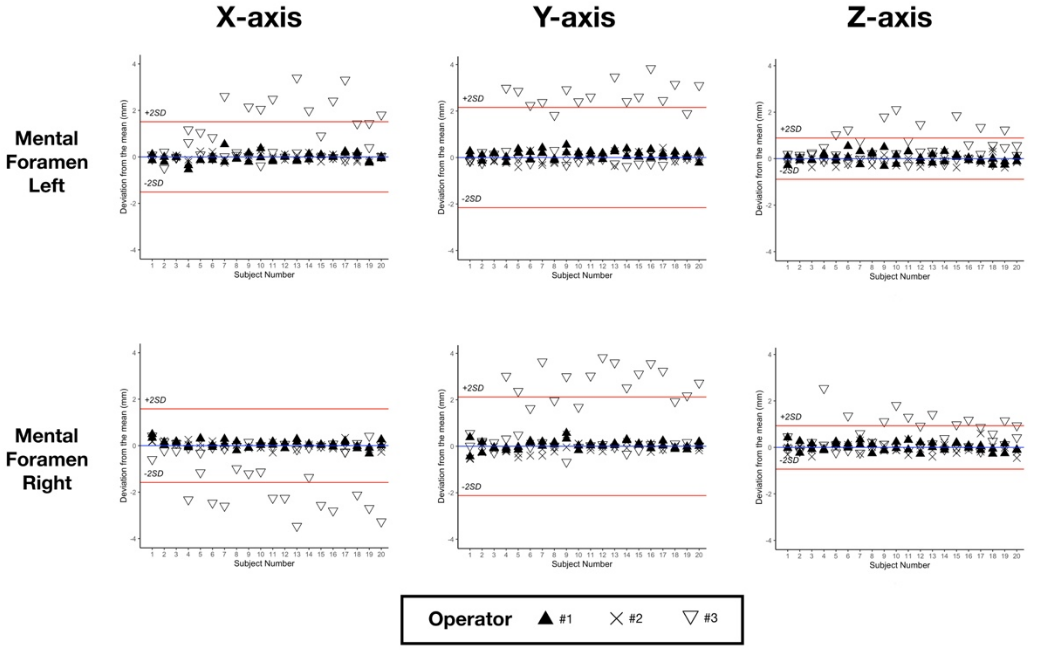


Figure SM1: Bland-Altman plots for mental foramen left and right with outliers, showing the deviations from the mean (blue line) of the 6 repetitions for the 20 subjects. Red lines show the ± 2*SD of reproducibility. SD, standard deviation.

Qualitative analysis of the results showed that the concerned landmarks had been localized in the most distal part of the mental foramina, contrary to the agreed upon definition which demanded placing these landmarks on the most mesial side of the foramen. Consistently with ISO 5725 recommendations, the corresponding annotations were treated as missing data. Figure SM2 shows Bland-Altmann graphs of the results after removal of the outliers.

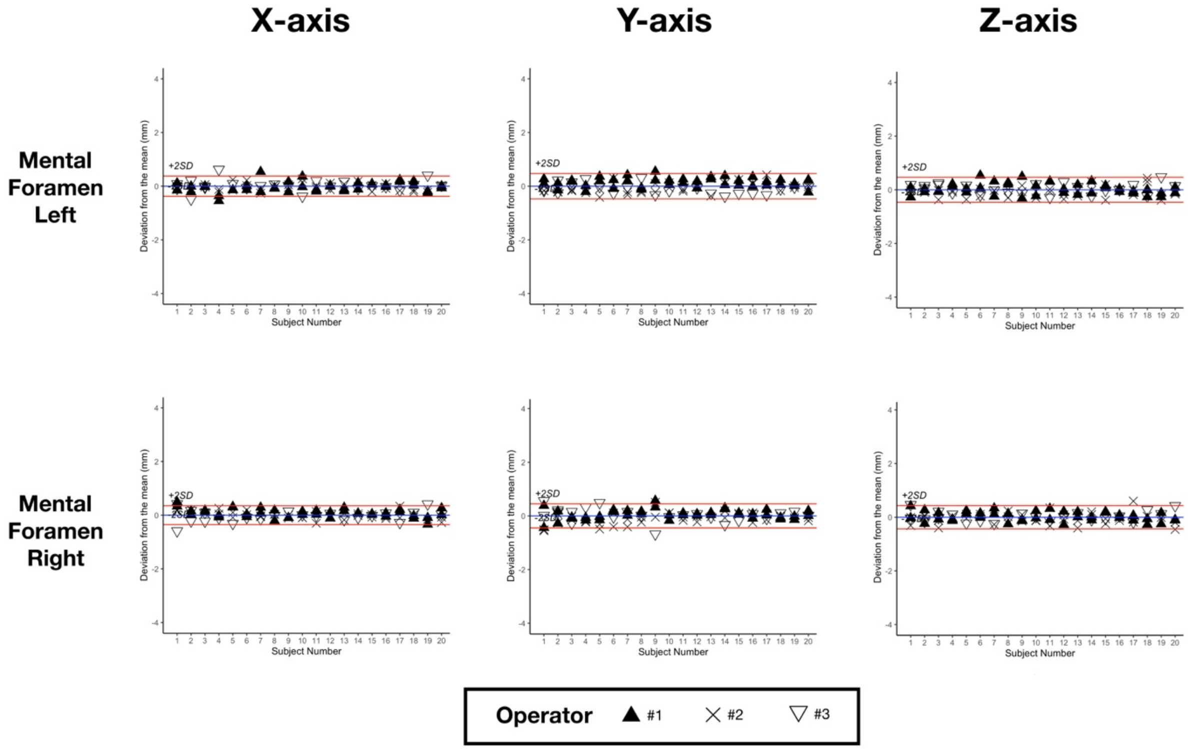


Figure SM2: Bland-Altman plots for mental foramen left and right without outliers, showing the deviations from the mean (blue line) of the 6 repetitions for the 20 subjects. Red lines show the ± 2*SD of reproducibility. SD, standard deviation.

**Supplementary Materials 3**: Angular distances between conventional and novel FH planes

**Method.** For each CT scan, four planes were computed using the means of all our operators' observations. Plane definition and labelling followed Pittayapat *et al.*'s publication: FH 1, FH 2, Plane 1 and Plane 3 (Table ST1). The absolute angular differences between each pair of planes were then computed, using trigonometry to calculate the angles between the normals to the planes.

| **Plane** | **Definition** |
|---|---|
| Frankfort horizontal plane 1 (FH 1) | FH by connecting mid-Po, Or-R and Or-L |
| Frankfort horizontal plane 2 (FH 2) | FH by connecting mid-Or, Po-R and Po-L |
| Plane 1 | A plane connecting mid-Or, IAF-R, IAF-L |
| Plane 3 | A plane connecting Or-R, Or-L and mid-IAF |

Table ST1: Definition of the 4 planes used in supplementary analysis

**Results.** Absolute angular differences between each pair of planes are summarized in Table ST2.

| Plane | Angular measurement (°) | |
|---|---|---|
| | Mean | SD |
| FH1 – FH2 | 0.98 | 0.57 |
| FH1 – Plane 1 | 2.94 | 1.34 |
| FH1 – Plane 3 | 2.04 | 1.34 |
| FH2 – Plane 1 | 2.59 | 1.27 |
| FH2 – Plane 3 | 2.41 | 1.21 |

Table ST2: Mean absolute angular differences and standard deviations between each pair of planes. FH, Frankfort horizontal plane; SD, standard deviation
